# A Framework for Advancing Sustainable MRI Access in Africa

**DOI:** 10.1101/2022.05.02.22274588

**Authors:** Udunna C Anazodo, Jinggang J Ng, Boaz Ehiogu, Johnes Obungoloch, Abiodun Fatade, Henk JMM Mutsaerts, Mario Forjaz Secca, Mamadou Diop, Abayomi Opadele, Daniel C Alexander, Michael Dada, Godwin Ogbole, Rita Nunes, Patricia Figueiredo, Matteo Figni, Benjamin Aribisala, Bamidele O Awojoyogbe, Christian Sprenger, Alausa Olakunle, Dominic Romeo, Francis Fezeu, Akintunde T Orunmuyi, Sairam Geethanath, Vikas Gulani, Edward Chege Nganga, Sola Adeleke, Ntusi Ntobeuko, Frank J Minja, Andrew G Webb, Iris Asllani, Farouk Dako, the Consortium for Advancement of MRI Education and Research in Africa (CAMERA)

## Abstract

MRI technology has profoundly transformed current healthcare and research systems globally. The rapidly growing burden of non-communicable diseases in Africa has underscored the importance of improving access to MRI equipment as well as training and research opportunities on the continent. The Consortium for Advancement of MRI Education & Research in Africa (CAMERA) is a network of African experts, global partners, and ISMRM/ESMRMB members implementing novel strategies to advance MRI access and research in Africa. To identify challenges to MRI usage and provide a framework for addressing MRI needs in the region, CAMERA conducted a Needs Assessment Survey (NAS) and a series of symposia at international MRI society meetings over a 2-year period. The 68-question NAS was distributed to MRI users in Africa and completed by 157 clinicians and scientists from across Sub-Saharan Africa (SSA). On average, the number of MRI scanners per million people remained at <1, of which, 39% were obsolete low-field systems yet still in use to meet clinical needs. The feasibility of coupling stable energy supplies from various sources has contributed to the growing number of higher-field (1.5T) MRI scanners in the region. However, these systems are underutilized with only 8% of facilities reporting clinical scans of 15 or more patients daily per scanner. The most frequently reported MRI scans were neurological and musculoskeletal. Our NAS combined with the World Health Organization and International Atomic Energy Agency data provides the most up-to-date data on MRI density in Africa and offers unique insight into Africa’s MRI needs. Reported gaps in training, maintenance, and research capacity indicate ongoing challenges in providing sustainable high-value MRI access in SSA. Findings from the NAS and focused discussions at ISMRM and ESMRMB provided the basis for the framework presented here for advancing MRI capacity in SSA.

**Graphical Abstract:** 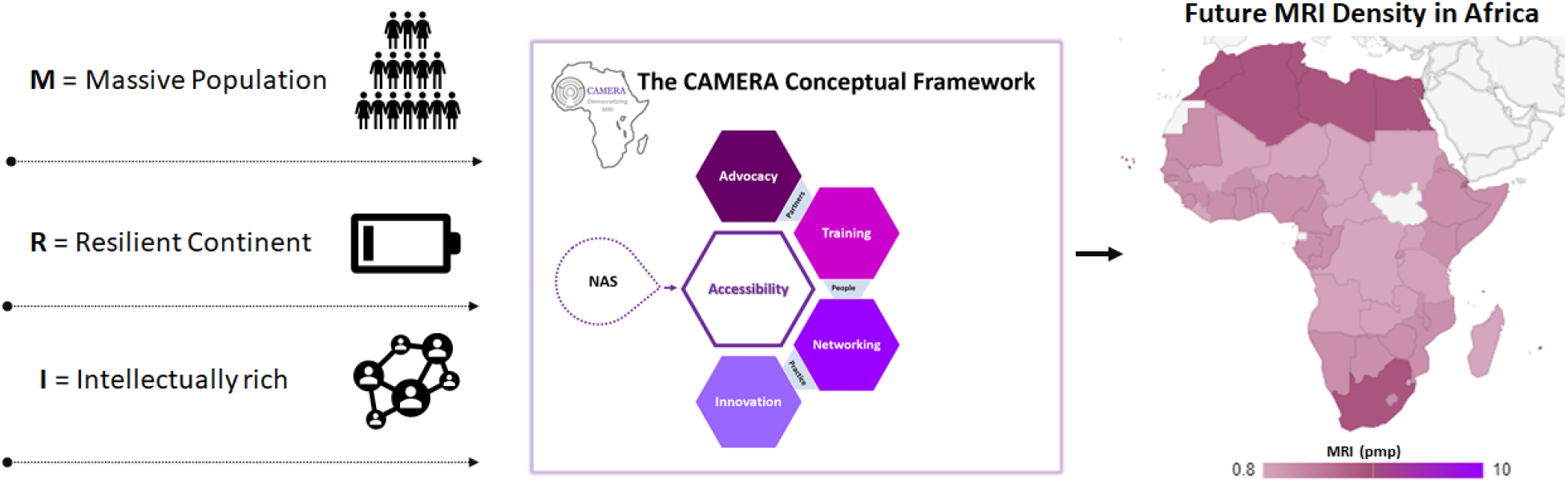

Africa has a massive population with few infrastructural resources and an untapped potential to effect transformative change in healthcare. To advance MRI access across all African countries and meet the sustainable development goals of improving health and wellbeing in low-resource settings over the next decade, the MRI community is called to partner with CAMERA to create enabling clinical and research MRI environments that will utilize the rich intellectual resources in Africa to realize lasting and equitable MRI for all Africans and the world at large.

## Introduction

The use of magnetic resonance imaging (MRI) as standard of care has grown significantly since its inception in the 1970s. This rapid growth is due in large part to the versatility of MRI in the management of non-communicable diseases (NCDs). Yet, despite a thriving overall trend in the proliferation of MRI, in regions with limited resources, such as Africa, MRI availability and use are restricted by high acquisition and maintenance costs; limited access to the infrastructure; lack of expertise required to run and maintain the equipment; and other region-specific factors^1,2^. In 2016, there were only 84 MRI units serving West Africa’s population of over 370 million, with over two thirds of these units located in Nigeria^2^. In 2017, the Africa Region had 0.7 MRI scanners per million people—the lowest density of MRI scanners among six regions defined by the WHO— followed by 1.1 in the South-East Asian Region, 2.8 in the Eastern Mediterranean Region, 4.1 in the Region of the Americas, 5.4 in the Asia Western Pacific Region and 11.6 in the Eurasian Region^3^. Given the centrality of MRI in the management of NCDs, increased availability and utilization of MRI services are critical to improving healthcare access and quality in Africa, in particular in Sub-Saharan Africa (SSA) where availability and access to MRI are even more restricted than in the rest of the continent.

Though NCDs are often associated with high-income countries, their burden in SSA is significant and growing due to the aging population, the drastically changing environment (e.g. air pollution)^4,5^, increased urbanization^6^, and changing behavioral trends such as tobacco use, sedentary lifestyles, excess alcohol intake and unhealthy diets^7^. From 1990 to 2017, while the disability-adjusted life-years (DALYs) due to NCDs decreased in high-income countries, in SSA, in contrast, the DALYs increased from 90.6 million to 151.3 million, corresponding to an increase in the proportion of DALYs due to NCDs from 18.6% to 29.8%^7,8^. Though historically, disease burden in SSA has been largely driven by communicable, maternal, neonatal, and nutritional (CMNN) diseases, the age-standardized DALY rate due to NCDs in 2017 was similar to that of CMNN diseases (21,757 per 100,000 population vs 26,491)^8^. Among NCDs, leading causes of disability in SSA are cardiovascular diseases (22.9 million DALYs), neoplasms (16.9 million), mental disorders (13.6 million) and digestive diseases (12.6 million)^8^. As NCDs related disabilities continue to increase in the region, it is critical to address barriers to NCD management in order to achieve the global target of reducing premature mortality from NCDs by one third by 2030^9^.

Increasingly, MRI use has expanded to take on an integral role in the management of NCDs, including management of cardiovascular disease, which is the leading cause of disability in SSA. While cardiovascular disease has decreased globally over the past decades^10^, the incidence is steadily increasing in SSA^11,12^. By enabling accurate assessment of cardiac morphology, function, flow, perfusion, acute tissue injury, and fibrosis, MRI can help determine disease etiology and risk, thus facilitating effective management of heart failure and other cardiomyopathies^13^ – even at low magnetic field strengths^14^.

MRI is the recommended standard of care in the diagnosis and treatment of neurological diseases, including but not limited to seizures^15^, strokes^16,17^ and dementia^18^. MRI can help determine seizure etiology in epilepsy management^15^, while in stroke imaging where recent evidence suggests that the incidence and prevalence of stroke in Africa has grown to 2-3 times that in Western Europe and the USA^19^, MRI can aid assessment of tissue perfusion, vascular flow, and treatment response in acute and subsequent phases of stroke^16^. Of note, the high prevalence of sickle cell disease in SSA makes neuroimaging particularly important due to the cerebrovascular implications of the disorder. Beyond NCDs, MRI can play a paramount role in the management of infectious diseases, such as HIV and malaria^20–22^.

In the context of a growing global cancer burden, access to imaging services can avert millions of cancer deaths annually worldwide^3^. MRI is a core component of cancer management in high income countries as it enables early and accurate diagnosis of cancer, thereby facilitating the treatment of neoplasms at a stage when they are more likely to be treatable. MRI is also used for assessing response to cancer therapy thus aiding physicians in making effective and economical treatment decisions^23^. Of note, MRI can reduce healthcare costs by preventing unnecessary procedures or tests^3^. While the direct benefits of MRI on cancer survival are difficult to quantify due to the complexity of cancer management, in Europe, cancer survival directly correlates with the number of MRI units per capita, supporting the view that MRI benefits cancer management^24^.

In a study published in *Lancet Oncology*, researchers estimated that a scale-up of imaging modalities (ultrasound, x-ray, CT, MRI, PET, and SPECT) in 2020-30 would avert 207,800 deaths in Africa, comprising 3% of total cancer deaths in the region and resulting in 4.64 million life-years saved^3^. Beyond humanitarian benefit, a scale-up of imaging modalities is also projected to yield significant economical returns. With an estimated cost of US$0.46 billion, a scale-up of imaging modalities in Africa from 2020-30 is projected to result in productivity gains of $27.38 billion with a net benefit of $26.93 billion^3^. In other words, there is an estimated return of $59.97 per $1 investment.

Despite the clear and urgent need to invest in medical imaging in SSA, a gap in knowledge and access to high value MRI services in the region persists. The aim of this study is to address this gap by: (1) Providing an assessment of access to high value MRI services in SSA using findings from the Needs Assessment Survey (NAS) developed and performed by the Consortium for Advancement of MRI Education and Research in Africa (CAMERA). (2) Describing the needs for MRI development in the region from recent discussions about the current state of MRI in SSA from CAMERA symposia at the International Society of Magnetic Resonance in Medicine (ISMRM) and European Society of Magnetic Resonance in Medicine and Biology (ESMRMB) is provided. (3) Presenting a framework informed by the NAS and symposia for addressing the need for MRI access and development in the SSA.

## Methods

CAMERA is a network of African experts, global partners, and ISMRM/ESMRMB members implementing disruptive approaches that are advancing MRI access in Africa through training and research capacity building. In May 2020, CAMERA initiated a Needs Assessment Survey (NAS) study which was designed as described below.

### Needs Assessment Survey

The NAS was designed to identify MRI needs unique to Africa based on a pilot field survey of seven MRI facilities in Nigeria and the RAD-AID™ Radiology-Readiness Survey^25^. A French and English version of the CAMERA NAS were created using Google Forms and distributed via email and WhatsApp messaging to radiologists, radiographers, and radiology organizations in Africa through the CAMERA member network. The participating radiology organizations were: Association of Radiologists in Nigeria (ARIN), Radiographers Registration Board of Nigeria (RRBN), Société de Radiologie d’Afrique Noire Francophone (SRANF) and Pan African Congress of Radiology and Imaging (PACORI).

The NAS form consisted of 68 questions divided into eight components aiming to capture key needs along 4 central themes/targets: 1) availability and access, 2) personnel training and education, 3) research translation and 4) sustainable technology (Table 1). The first 8 components consisted of close-ended questions to provide insight on: 1) facility information, 2) scanner description, 3) maintenance, 4) usage and indications, 5) facility infrastructure, 6) personnel training and education, 7) health economics, and 8) research capacity. Optional short answer open-ended questions were included at the end of the survey to allow responders to provide additional context to MRI needs as well as suggestions for potential solutions.

**Table 1:** CAMERA NAS Structure

| Challenge Theme/Target | NAS Component |
| --- | --- |
| 1. <u>Availability and Access</u><br>. Density (number of MRI scanners or personnel/facility)<br>a. Location (public/private/teaching - rural vs urban)<br>b. Field strength (low/high)<br>c. Utility - Indications, how well the system is utilized | Facility information<br>Scanner description<br>Usage and indications |
| 2. <u>Personnel Training</u><br>. Capacity to utilize (image acquisition) – acquire good images and run the system well (radiographer)<br>a. Capacity to utilize (image interpretation) – diagnose and provide outcomes (Radiologist)<br>b. Access to CME education and workshops | Personnel training and education |
| 3. <u>Research Translation</u><br>. Capacity to drive MRI innovations to solve local health care needs<br>a. Capacity for use of MRI in clinical trials and large population studies<br>b. Relevant imaging technology development (hardware and software)<br>c. Networking and access to research networks | Facility information<br>Facility infrastructure<br>Scanner description<br>Research capacity |
| 4. <u>Sustainability</u><br>. Maintenance and servicing access and frequency<br>a. Life cycle of MRI scanner<br>b. Power supply infrastructure<br>c. Funding to sustain the MRI facility and scanner. | Facility infrastructure<br>Scanner description<br>Maintenance<br>Health economics |

To meet one of CAMERA’s mission of strengthening MRI networks within Africa, participants were asked to opt-in to be notified and included in networking efforts by providing their center and primary contact information. This optional input is only accessible to The CAMERA Environmental Scan Task Force, as disclosed on the survey, and is not reported in this study. The English version of the CAMERA NAS is appended in the supplementary materials (Table S1).

To assess the MRI density in the region, the NAS data were supplemented with data on the total number of MRI scanners per country from the World Health Organization (WHO) 2010-2014 Global Health Observatory data repository^26^ and the International Atomic Energy Agency (IAEA) IMAGINE Database^27^. Both databases were accessed in August 2020, with the most recent (IMAGINE) data information retained for data analysis. Population density was obtained from World Bank estimates. The MRI density for each African country was calculated as the number of MRI machines per million inhabitants.

To further explore research capacity across the region, the total number of MRI papers published between 2018 and 2020 were tabulated from the Journal Citation Reports Journal Profile page. Ten Journals, listed in the Radiology, Nuclear Medicine & Medical Imaging and the Spectroscopy, Physics, Atomic & Molecular categories of the Journal Citation Reports, and whose scope falls predominantly within MRI research were included^28^. To gain a better understanding of the research performance in the region, data from all regions of the world captured for each journal were included in this analysis and classified based on The World Bank Country Income Groups^28^.

### CAMERA Societies Meetings: Agenda

Between 2020 and 2021, CAMERA held three symposia at ESMRMB and ISMRM. The goal was to highlight Africa’s complex and unique MRI needs and outline opportunities for breakthrough increase to MRI access and research. These symposia expanded on concepts central to the 4 themes captured in the NAS: availability and access, personnel training and education, research translation, and sustainable technology.

Briefly, the meetings were organized through proposal submissions to the ESMRMB congress planning committee and the ISMRM virtual meeting approval process. Speakers and moderators were sourced from the network of MRI users and experts working in the region. All meetings were held virtually using the standard Zoom video conferencing platform, except for the symposium at the 2021 ESMRMB congress, which was facilitated by Conventus Congress Management & Marketing GmBH (Jena, Germany). The 2020 ESMRMB Congress symposium and the 2021 ISMRM Virtual Meeting were free to attend, while the 2021 ESMRMB Congress symposium was limited to attendees who registered to attend the entire Congress. The symposia featured 2 - 5 talks ranging from 20 - 30 minutes per speaker, followed by a 1-hour panel discussion to draw more inputs from the attendees (Table 2).

**Table 2:** Agenda for The CAMERA Symposia

|  | Topic title | Speaker Name and Affiliation |
| --- | --- | --- |
| ***ESMRMB 2020 Congress, October 1 2020*** |  |  |
| 1 | Doing much with little: benefits and challenges of low field MRI in Africa | Godwin Ogoble MD, University College Hospital, University of Ibadan, Nigeria |
| 2 | Expanding the imaging enterprise and implementation of artificial intelligence in Africa | Farouk Dako MD, Perelman School of Medicine, University of Pennsylvania |
| 3 | Installation of an MRI system in Mozambique – implementation, difficulties, and impact | Mario Forjaz Secca PhD, Central Hospital of Maputo, Maputo, Mozambique |
| ***ISMRM Virtual Meeting: ISMRM Spotlights Africa, February 25 2021*** |  |  |
| 1 | MRI in practice in Africa | Abiodun Fatade, MBBS, Crestview Radiology, Lagos, Nigeria |
| 2 | Diagnosing cardiac disease using MRI in Sub-Saharan Africa: Battling Cardiac disease in a perfect storm | Ntobeko Ntusi, MBChB, Department of Medicine, University of Cape Town, South Africa |
| 3 | Breast MRI in Africa: Current status and challenges of implementation in screening, diagnosis, staging and follow-up | Nagla Abdel Razek, MD, Department of Radiology, Cairo University, Egypt. |
| 4 | The role of machine learning in MRI in low-and middle-income countries | Daniel Alexander, PhD University College London, UK |
| 5 | Developing techniques for low-field MRI applications in Sub-Saharan Africa | Johnes Obungoloch, PhD, Mbarara University of Science and Technology, Uganda |
| ***ESMRMB 2021 Congress, October 8 2021*** |  |  |
| 1 | Reimagining accessible MRI education in Africa: Physics + Medicine Your Tube | Abayomi Opadele MSc, Hokkaido University, Japan |
| 2 | MRI in clinical practice in Africa – A radiographer’s perspective | Harrison Aduluwa BSc, MPH, Euracare Multi-Specialist Hospital, Nigeria |

## Results

### Outcome of the CAMERA Needs Assessment Survey

The NAS was conducted from May to October 2020. A total of 172 responses were received from the following 12 SSA countries: Cameroon, Gabon, Gambia, Ghana, Kenya, Malawi, Niger, Nigeria, Somalia, South Africa, Tanzania, and Uganda. Out of all responses, 15 were discarded as duplicate entries by multiple individuals from the same institution. Results from the 157 CAMERA NAS responses are summarized in Figures 1–3. Results for each of the NAS survey entries – Availability and Access, Training and Education, Research Translation, and Technology Sustainability – are provided below.

**Figure 1:**
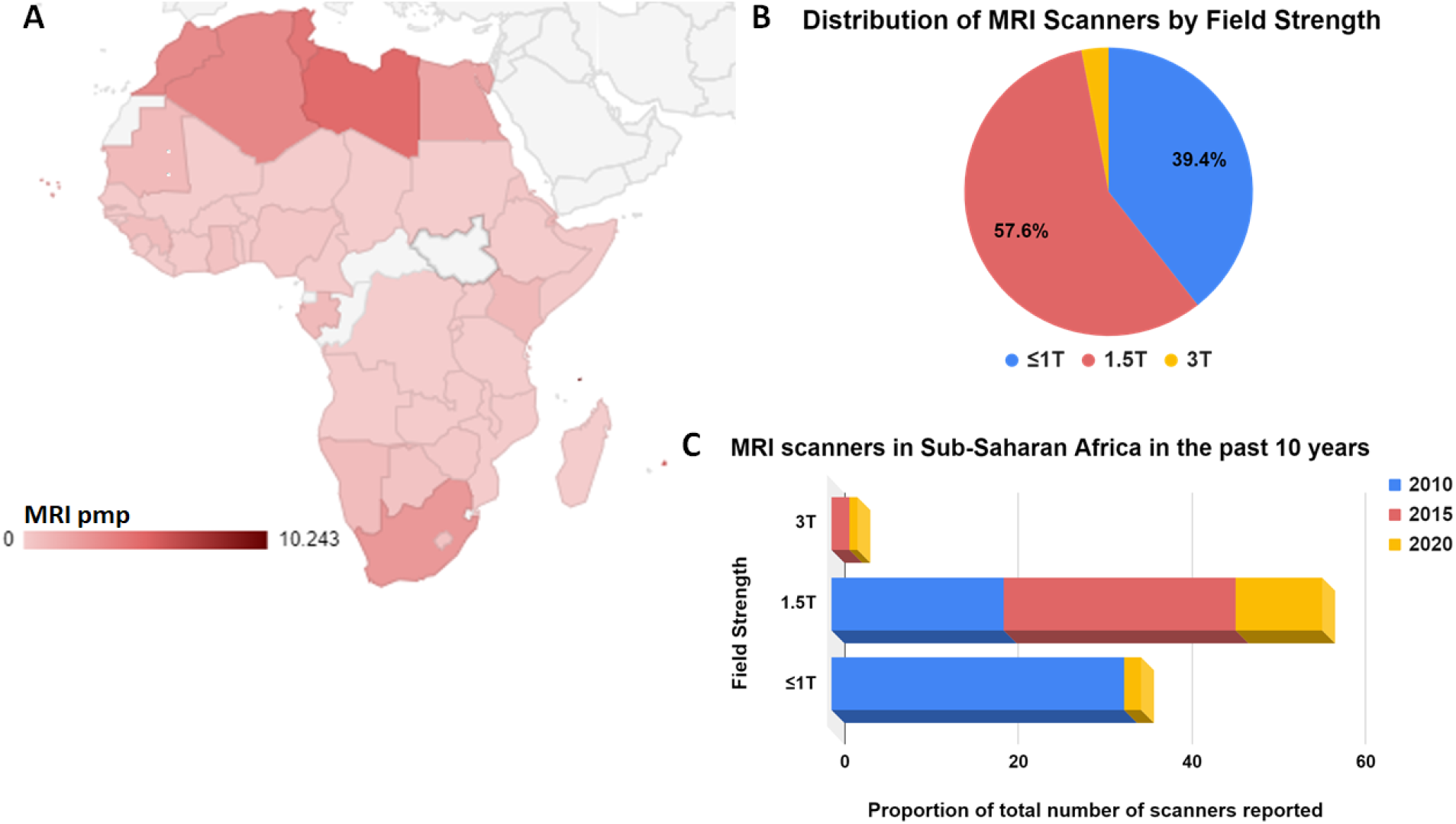
MRI Density and Proportion of Scanners by Field Strength. (**A**) Density of MRI across Africa demonstrating the number of scanners per million people (pmp) for each country with data (colored). (**B**) Distribution of MRI scanners in Sub-Saharan Africa by field strength show considerable proportion of reporting centers use low-field (≤1T), however there is a growing number of high-field (1.5T and 3T) scanners in the region but also ongoing procurement of low-field MRI (**C**)

#### Availability and Access

The combined data from CAMERA NAS, WHO^26^, and IAEA^27^ databases showed that MRI scanner density in Africa on average was 0.8 scanners per million people (pmp) (Figure 1A). Density data were unavailable for 5 of the 54 countries. Of the 49 countries that reported density data, 11 countries reported zero MRI scanners, a list that included highly populated countries such as the Democratic Republic of Congo with ∼95 million inhabitants, Niger with ∼26 million, and Mali with ∼21 million (supplementary materials Table S2). In contrast, Romania, with a population similar to Mali’s and one of the countries with the lowest gross national income in Europe, has 7-10 MRI pmp^27^.

Of the 157 NAS responses, 45% (70) reported not having an MRI scanner in their center. Of all the imaging departments/centers with MRI: 56.6% were public, including government funded academic healthcare centers (7.54%) and regional/tertiary and community hospitals (13.3%). The rest (43.4%) were private practices, which included only 2 public-private partnerships.

In total 110 MRI scanners were reported across the 87 centers, with 83 scanners reported as functioning at the time of the survey. Four of the 110 scanners were 3T, 58 were 1.5T, one 1T and 41 <1T. Three participants reported the total number of MRI scanners at their center but did not provide the field strength. The distribution of MRI by field strength is shown in Figure 1B. Three centers, one in Nigeria, one in Tanzania and one in South Africa, all affiliated with public universities reported working with 3T and 1.5T scanners at their center, with two of the four 3T scanners in the South Africa center. The single 1T reported was in a National Hospital in Nigeria. Out of the 42 respondents who reported working using low-field (≤1T) MRI in regular clinical practice, 35 worked in public/government centers. Notably, about 81% of the low-field systems were reported as having exceeded their obsolescence period (Figure 1B). However, a steady growth in the number of high-field scanners, primarily 1.5T, was also reported (Figure 1B).

#### Personnel Training and Education

All centers with an MRI reported having at least 1 radiologist and 1 radiographer on staff. The distribution of staff by discipline is shown in supplementary materials Table S3. However, 65% (57/87) of respondents reported not having a staff MRI physicist.

Figure 2 shows the proportion of radiologists and radiographers who regularly attend continuing education (CME) MRI events, their levels of participation (local/national/regional/international) and the common mode of participation/attendance (online/in-person). All 69 respondents who completed the section of the survey on interest in attending MRI workshops said their staff would benefit from attending local/national/regional MRI workshops. Out of 67 respondents, 63 responded ‘yes’ or provided further elaboration to the question related to perceived training opportunity challenge, while only 4 indicated no perceived challenge to training. Training cost, limited time off to attend training, and lack of available training courses/workshops/opportunities were commonly expressed as barriers to continuing education. Below, are few examples of the respondents’ unedited replies to training opportunity challenges:

**Figure 2:**
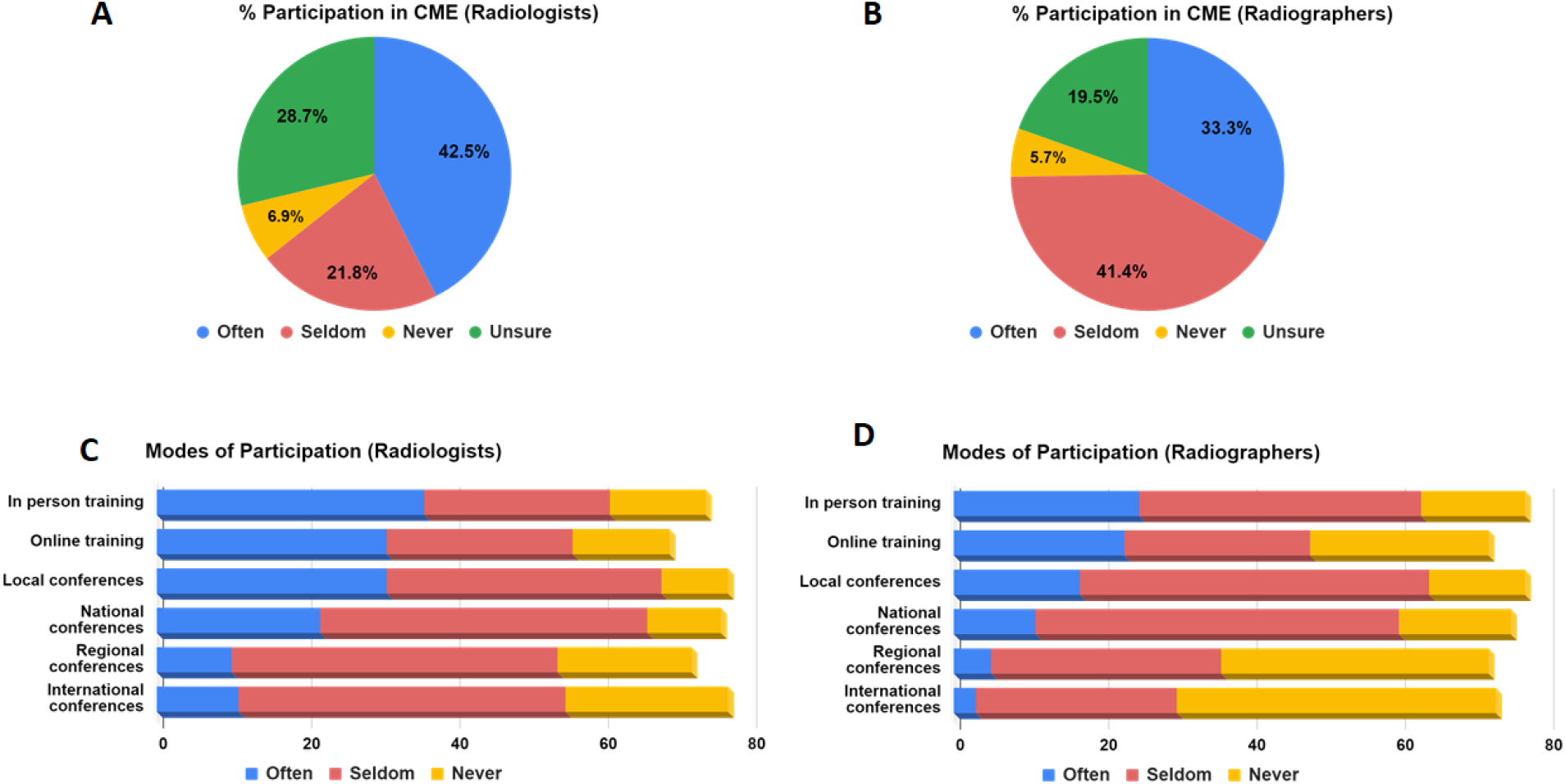
MRI Training and Education Access in SSA. Proportion (%) of CAMERA NAS respondents (n=87) who have participated in MRI meetings, workshops or conferences in the past 5 years often (2-3 times per year), or seldom (1 time per year) or never (**A** and **B**) over this period using various modes of participation (online or in-person) and at various levels of participation, locally (within their institution), nationally (within their country), regionally (within Africa) or internationally (abroad) (**C** and **D**).

> *“Given the challenges faced in my locality, there is increased in number of patient. Revolving technological advancement needs continuous training to meet the needs of the community I’m serving and impact the knowledge into my students in the University. Thank you”*
>
> *“We have very few Radiographers and the absence of 1 will increase TAT, cost and lack of sponsorship and total apathy to personnel development by management*.*”*
>
> *“We are motivated if we are not limited by costs of training*.*” “Training in English is a problem”*

#### Research Translation Capacity

Of the 87 MRI centers who completed the NAS, 52 (60%) lead or participate in research using MRI but only 63 (72%) have a research ethics board. Brain imaging applications, neuroscience, and musculoskeletal imaging applications were the most commonly indicated areas of MRI interest (Supplementary material, Figure S1).

Five of the ten MRI journals profiled on the Journal Citation Reports were official publications of MRI societies, with 2020 Impact Factor ranging from 2.31 to 5.36. In total, 83 countries published 8131 MRI-related papers between 2018 and 2020. Of these, 11 countries were from Africa and 7 from SSA. The total number of MRI publications from the region were 40 (Africa) and 17 (SSA), representing 0.5% and 0.2%, respectively, of the total MRI publications in the most recent 3 years (Figure 3, supplementary materials Table S4). In contrast, Upper Middle-Income regions such as Brazil, China, Turkey, and Iran had a combined 57 times more MRI publications compared to SSA, while High Income countries such as Canada, Chile, Germany, England, South Korea and the US published a combined 86.7% of the total MRI publications over the same period, representing 433 times the number of papers from SSA (Figure 3B, Figure S2, and Table S4). In SSA, Nigeria, Uganda, and South Africa were the only countries that published a total of 4 MRI research studies in the ISMRM (JMRI), ESMRMB (MAGMA) and Society of Cardiovascular Magnetic Resonance (SCMR) journals.

**Figure 3:**
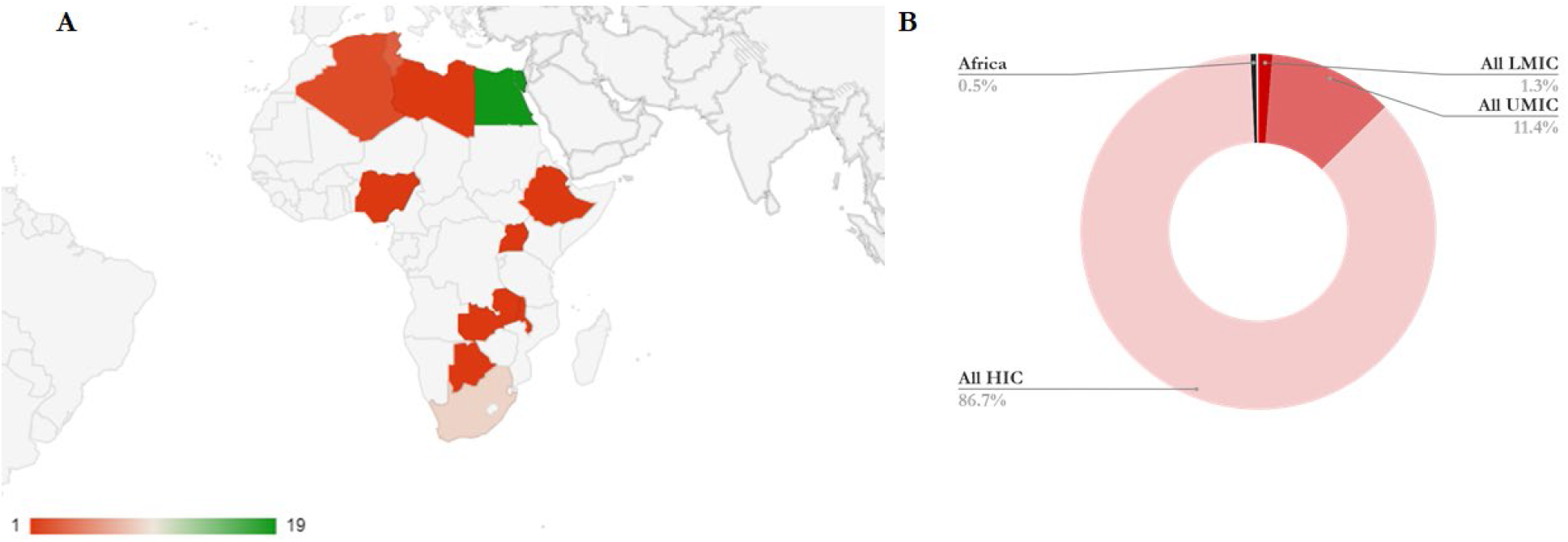
Research output. Total number of MRI publications in MRI exclusive journals between 2018 and 2020 for each country with data (colored). (**A**). All SSA countries with data minus South Africa (9) produced just a single publication each. (**B**) Africa as a whole produced ∼0.5% of the total MRI publications over this period in the 10 journals.

#### Sustainable Technology

In terms of utility, about 48% of centers perform clinical scans on ≤5 patients/scanner/day and only 8% scan ≥15 patients/scanner/day (Figure 4A). The top three reported clinical indications were brain, spine, and musculoskeletal (Figure S3). The average reported cost of an MRI examination was US$200. Payment for MRI services at the facilities were overwhelmingly from private or out-of-pocket sources (37% sole source or 89% as one of the sources of payment) compared to partial or full public funding including national insurance schemes (3.4% sole source or 35.6% as one of payment sources) or private insurance (0% sole source or 37.93% as one of payment sources), as shown in Figure S4.

**Figure 4:**
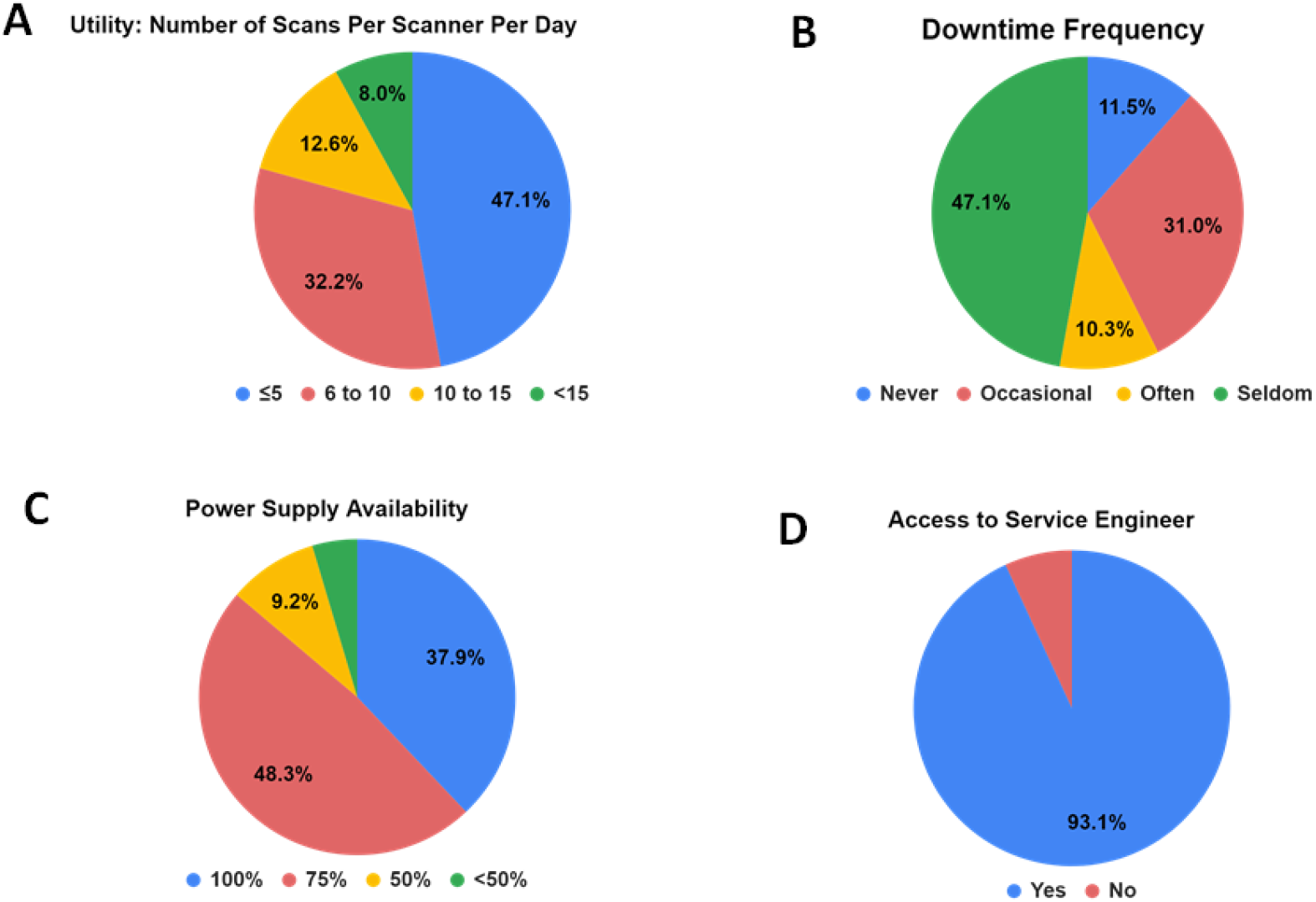
Sustaining high-value MRI in Sub-Saharan Africa. The proportion (%) of CAMERA NAS respondents (n=87) who have at least one MRI scanner reported the number of scans per scanner per day (**A**), their downtime frequency (**B**) (as never/none, occasional [1-3 times per month], often [1-2 times per week] or seldom [<1 times in a 6 month span]), how often they have steady power supply (**C**), and if they have access to regular service and maintenance support.

Of the 87 MRI facilities, 81 have access to a service engineer (Figure 4), as a staff/faculty member (5/81), third party (61/81), both (5/81) or through other means (5/81). The reported frequency of scanner maintenance/service from 81 respondents was 13% (11/81) for monthly, 34.5% (28/81) for 2-3 times per year, 11% (9/81) for annually, and 40.74% (33/81) for never. Downtime frequency and power supply availability reported by the 87 centers with MRI are shown in Figure 4.

Regarding infrastructure, up to 86% of facilities reported access to constant power supply (≥75% of 24hours/7days/week), 57.4% reported downtime frequency of at least 1-2 times/ week (often) or 1-3 times per month (occasional) (Figure 4B-C), 94% (82/87) reported access to a backup power supply, 83% (82/87) reported access to a picture archiving system (PACS) at their center, and 29% (25/87) use teleradiology (reading and interpretation at another facility).

### Outcome of the CAMERA Symposia at the MRI Societies Meetings

A total of approximately 120, 100, and 21 individuals attended the 2020 ESMRMB symposium, 2021 ISMRM virtual meeting, and 2021 ESMRMB symposium, respectively. The low attendance for the 2021 ESMRMB symposium was due to the relatively high registration cost (90 euros). On average 30% of attendees were resident in Africa.

Highlights of discussions from the three symposia are outlined in supplementary material Table S5. The discussions expanded on the issues of availability and access, personnel training and education, research translation, and sustainable technology. For each issue, challenges and opportunities for CAMERA and the MRI community were identified and discussed (Table S5). Major barriers that contribute to the ongoing low use of MRI in Africa were recognized as either infrastructural or institutional, both involving issues of availability and access, challenges in education and training, lack of research environment, high operational cost, and limited specialized expertise needed to sustain MRI technology (Figure 5). These challenges along with recommended solutions are briefly summarized below.

**Figure 5:**
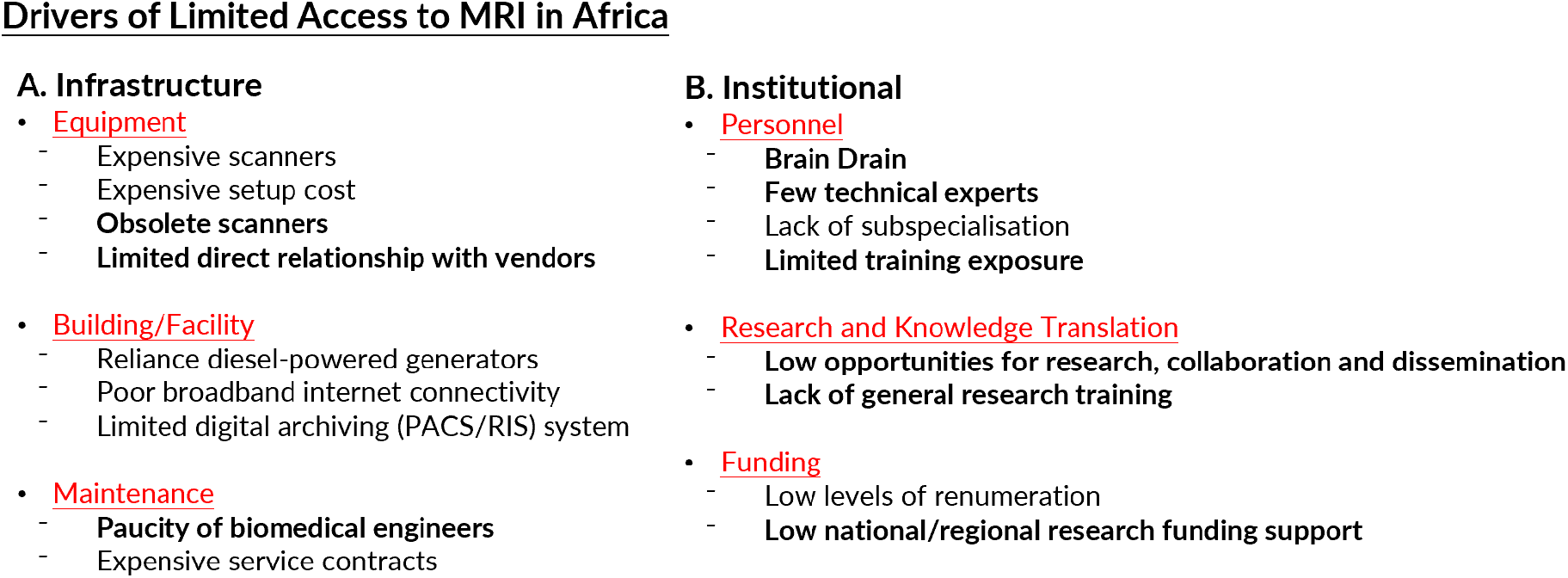
Barriers to high value MRI in Africa.

#### Challenges in Availability and Access to MRI

##### Acquisition costs

African countries are required to outsource MRI equipment as medical devices are rarely produced on the continent. As a result, the MRI equipment can be more expensive in Africa than in the West and other regions with locally sourced medical equipment. The lackluster medical manufacturing market not only hinders access to medical technology, but also denies Africa the technical expertise and skills development necessary for a sustainable healthcare industry.

##### Limited access to financial capital to support investment in MRI equipment

Due to poor health insurance coverage in the region, healthcare providers in SSA cannot generate adequate returns to justify investment in high-cost MRI equipment. Furthermore, healthcare providers have limited options for funding: financial companies are not healthcare friendly as medical equipment often cannot generate quick returns, while banks do not have existing lending schemes for medical devices. Small and medium healthcare providers are particularly disadvantaged in accessing loans due to their lack of banking history and limited collateral.

#### Challenges in Sustainable Technology

##### Scarcity of human capital to run, utilize, and maintain MRI technology

The limited experience of radiographers, radiologists, and biomedical engineers contribute to the general low throughput. Poor maintenance often renders MRI systems unusable far in advance of the expected product lifetime. In Uganda, $1.3 million USD worth of medical equipment was found to be non-functional across 17 medical facilities within two years of donation. In some cases, the equipment had never even been installed. Because a significant portion of medical devices in SSA is donated, recipient facilities often have limited decision-making power in what equipment they receive, which can result in a mismatch between a donor’s agenda and a facility’s needs. Recipient facilities may also lack resources and expertise to appropriately maintain and utilize the donated equipment. Furthermore, Africa’s brain drain exacerbates the shortage of medical professionals and strips the region of its best human resources^29,30^.

##### Poor electricity and technology infrastructure impedes effective MRI technology utilization

Limited access to stable electricity infrastructure prevents utilization of high-field MRI systems that require cryogens. Additionally, electricity from the power grid may be unclean and thus may damage expensive equipment^31^. The poor penetration of the Internet, computers and other technology presents challenges in healthcare management, including accessing and storing patient information, imaging files and referrals^32^.

#### Proposed Solutions

##### Invest in low-field MRI as a cost-effective and adaptable alternative to high-field MRI

Low-field MRI systems are significantly cheaper than high-field systems and not only does it have the potential to address most clinical needs in SSA, but also has manifold advantages over high-field MRI^14,16,33^. Furthermore, low-field equipment requires less technical expertise to run and maintain. Important, low-field equipment is significantly less heavy and easier to transport than high-field systems thus reducing transportation costs and challenges associated with setting up the equipment. Since low-field systems are more robust and can better handle fluctuations in electricity, they are more suited to low-resource settings in SSA where power outages can occur frequently^34^. Despite breakthrough technological advancements in low-field MRI, the image quality is relatively low when compared to those from high-field. However, mounting evidence suggests that low-field MRI image quality can approach that of high-field MRI, especially for core clinical applications^34–36^.

##### Apply machine learning technology to enhance healthcare services and delivery

One of the ways to boost the quality of the low-field MR images is using Image Quality Transfer (IQT) ^35^approaches that uses Artificial Intelligence (AI) to propagate information from expensive high-quality images acquired at high-field to the inexpensive, low-quality images such as those acquired with low-field. Beyond image enhancement, AI technology can enhance healthcare delivery by enabling faster diagnoses as well as facilitating patient referrals. Though the role of AI technology continues to grow in the medical field, there is a lack of data diversity especially from low- and middle-income countries^37^. Countries in SSA need to develop legal and regulatory frameworks to support data science research that may pave the way for precision medicine and other medical advances.

##### Digitize radiology and adopt enterprise imaging to improve data management

Digitizing radiology is a critical step for medical facilities as it enables teleradiology services, inter-institutional image exchange, robust storage of imaging data, as well as cost and time savings for patients. Furthermore, digitizing radiology is essential to the adoption of enterprise imaging, which involves integrating all healthcare data into one system that can be accessed using a universal viewer^32^. Instead of storing patient information and imaging files in disparate departmental systems, with enterprise imaging, all data is stored in a central database. The benefits of enterprise imaging include improved patient safety, a reduction in redundant tests, and remote access to imaging on mobile devices^32^.

##### Create vendor financing programs

Original Equipment Manufacturers (OEMs) and equipment vendors can create vendor financing programs to facilitate healthcare providers in obtaining loans for medical equipment. In 2019, GE Healthcare and the Medical Credit Fund entered into a partnership to improve access to medical equipment in Kenya^38^. Under this partnership, small and medium private healthcare providers can borrow up to $100,000 with a 24-months repayment program to purchase GE Healthcare manufactured medical equipment^38^. Similar partnerships in other countries in SSA can allow small and medium healthcare providers to secure loans to purchase MRI systems.

##### Establish skills development and training programs for MRI personnel

OEMs, ISMRM and MRI networks such as CAMERA should regularly host skills development and training programs for radiologists, radiographers, and biomedical engineers in SSA. These programs can produce a more skilled local workforce for medical facilities to rely on for the running and maintenance of MRI systems. Further, an increased number of medical professionals can help to offset the effects of Africa’s brain drain. Dr. Naglaa Abdel Razek, who now runs a breast MRI program in Egypt after partaking in a European Society of Breast Imaging (EUSOBI) training program, is a prime example of how investing in human resources can generate remarkable returns for local populations. Another striking example is installation of new Siemens Healtineers magnets by Mr. Alausa Olakunle, a local third-party biomedical engineer who in 2015 received skills training on the Siemens platform at Cary, USA to enable installation of OEM magnets across Africa, including recent installations of a 3T MRI at the Ruby Medical Center in Kampla, Uganda and the 1^st^ 0.55T MRI (MAGNETOM Free.max, Siemens Healthineers, Erlangen, Germany) in Africa, at Hospital Geral De Cabinda, in Cabinda, Angola.

##### Advocate for increased public and private investment in medical imaging research

Locally sourced medical equipment and consumables will provide significant cost reductions for both the healthcare provider and the patient. Increased production of medical research in SSA will facilitate knowledge transfer of best clinical practices in low resource settings.

##### Couple energy sources to provide stable electricity solutions for healthcare providers

In the absence of stable electricity infrastructure, healthcare facilities can couple up different power sources to provide a reliable source of electricity for MRI equipment and other medical devices. For example, Crestview Radiology Limited, a busy private clinic in Nigeria with a 1.5T MRI (20-25 MRI scans/day) and 1 CT (10-15 scans/day), developed a local solution to clean uninterrupted power supply by coupling the electricity provided from the National Grid and alternative power sources to support full operation of their imaging devices (Figure 6, Figure S5). To ensure that only clean power is supplied to their imaging scanners, the clinic connects their imaging equipment to the power supply via automated voltage regulators/UPS (uninterruptible power supply) systems, while maintaining two backup power diesel generators. The backup generators, most of the time, serve as the main source of power, running at alternate times with one serving as a backup to the other. To ensure proper operations and minimize downtime, the clinic has an Information Technology (IT) personnel that regularly monitors the performance of the generators and the automatic voltage regulators/UPS systems.

**Figure 6:**
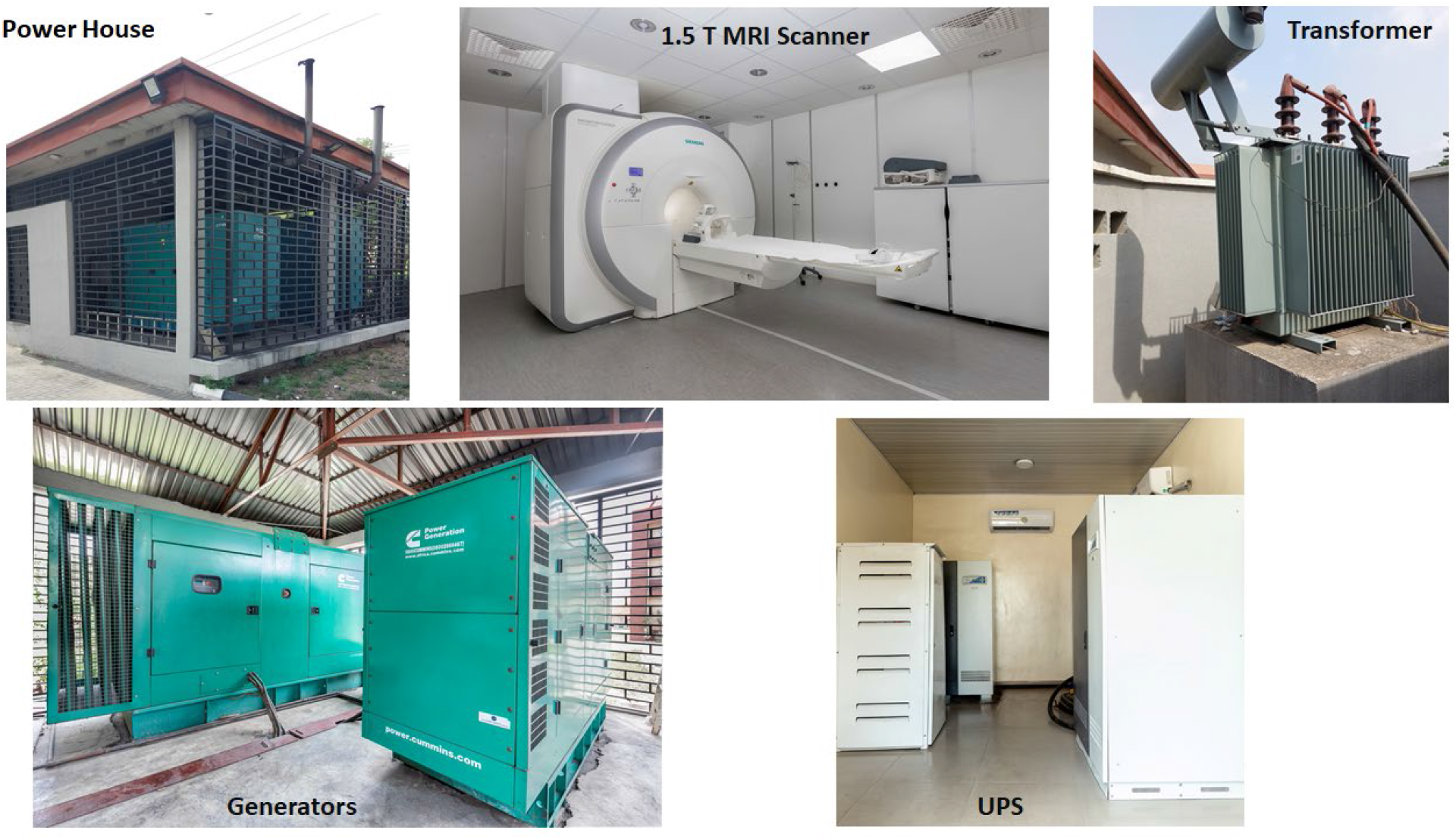
Energy Solutions for Steady Power Supply in SSA Medical Imaging Facilities. Images of the local energy solution at Crestview Radiology Limited in Lagos, Nigeria for steady power supply for operation of the MRI equipment and other imaging devices.

### The CAMERA Framework

CAMERA was established in 2019 as an ad hoc working group of the ESMRMB in direct recognition of these crucial challenges. Since then, CAMERA has formalized its goals and strategic plan via a NAS and consultations with several investigators and academic stakeholders through the ISMRM and ESMRMB engagements.

#### Vision

To make MRI accessible to provide high value care in Africa and to advance innovation geared toward solving the relevant healthcare needs of the continent.

#### Mission

1. **C**reate enabling clinical and research MRI environments
2. **A**dvance MRI education – train and retain
3. **M**aintain, expand, and strengthen MRI networks within Africa

1. **E**mpower African researchers toward need-specific innovation
2. **R**ealize lasting partnerships with academia, industry, and relevant organizations
3. **A**dvocate for diversity and inclusion in MRI

#### Structure

The structure of CAMERA is outlined in Figure S6 of the supplementary material. The governance structure provides strategic direction for the day-to-day activities of CAMERA, ensuring that effective oversight and adequate representation of African MRI clinicians and scientists is balanced with global partners and stakeholder engagements. Five task forces are being established to address the specific MRI needs identified in this study.

#### Strategy and Approach

We understand that in SSA, training builds on access to technology, and research excellence depends on high-capacity personnel and sustainability is required for Africa to thrive in relevant MRI research translation and clinical application. In this regard, to increase high value MRI use in Africa, CAMERA will focus activities on four core pillars (Figure 7):

**Figure 7:**
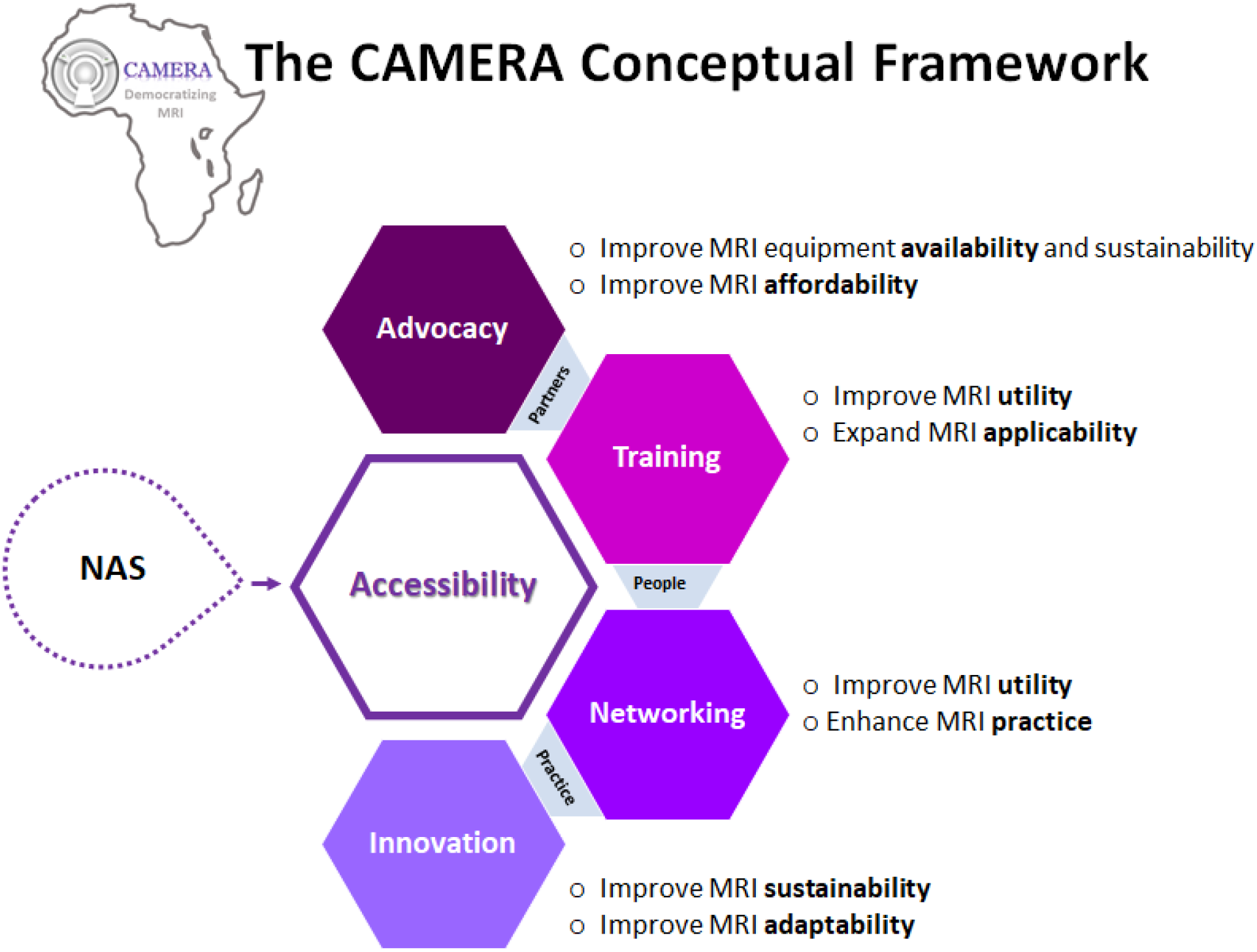
The CAMERA Conceptual Framework for Advancing MRI Access in Africa. The CAMERA Conceptual Framework integrates four core pillars of activities (advocacy, training, networking and translational research innovation) to address Africa’s multifaceted MRI accessibility challenges and transform healthcare across Africa. The framework is informed by a Needs Assessment Survey (NAS) and series of focused meetings where accessibility gaps were identified. This framework outlines an approach for working with partners to advocate for affordable MRI technology which will provide enabling environments for qualified people to use best practices to drive and democratize MRI across Africa.

##### 1. Advocacy

a. Raise awareness on the role of MRI in management of Africa’s unique healthcare needs through webinars and symposia.
b. Facilitate partnerships and collaboration between MRI OEMs, distributors and users in the region to enhance procurement, effective use, and maintenance of scanners.
c. Advocate for funding support for MRI research including from non-traditional sources (e.g., the recent award from Chan Zuckerberg Initiative (CZI)).

##### 2. Effective Training and Mentorship

a. Use a train-the-teacher approach with tiered learning competency to upskill and mentor an interdisciplinary group of emerging influential MRI experts in targeted areas (i.e., brain imaging and low field MRI) that will lead to: i) high value acquisition and workflow (i.e., protocol optimization, pulse sequence programming, MRI safety, hardware development, and enterprise imaging), ii) high image quality (i.e., image processing and analysis and imaging informatics), iii) high value diagnostics (i.e., case-based learning approach), and iv) improved research translation capacity (i.e., grant writing, publication, research ethics, data management).
b. Use a decentralized training approach to enable wide replicability, particularly to non-Anglophone regions. Novel training modes and tools such as the use of social media (e.g. Abayomi Opadele’s Physics+Medicine Youtube series), podcasting, and twitter will be employed to asynchronously provide educational opportunities.

##### 3. Networking

a. Provide access to regular cross-regional meetings to foster regional collaboration, support local isolated groups, and expand the use of MRI in for example cardiac application, learning from regional leaders such as Professor Ntobeko Ntusi’s group at University of Cape Town.
b. Provide access to global MRI collaboration through establishment of the African Chapter of the ISMRM as well as partnerships with other international imaging societies (ESMRMB, SCMR, MICCAI etc.)
c. Develop a platform for MRI resources and user-forums to share best practices, especially given that most of the scanners in SSA, including the 1.5 and 3 T systems are reconfigured versions and SSA users do not have direct connection to OEMs.

##### 4. Translational MRI Technology Innovations

a. Develop, support, and implement relevant MRI innovations that will facilitate sustainable MRI technology through: i) AI-enabled retooling of existing MRI scanners, ii) clinical application of contrast-free imaging techniques to provide no-cost solutions to consumables, such as the use of arterial spin labeling (ASL), iii) ease-of-use or graphical-user-interface (GUI) open-sourced image processing and analysis pipelines.

## Discussion

We examined availability and use of MRI in Sub-Saharan Africa to provide a novel framework to address MRI needs. Over a two-year period we identified threats to access, use, and development of MRI in the region from a needs assessment survey and series of focused discussions at ISMRM and ESMRMB. We observed a promising increase in high-field MRI despite limited power supply, physical infrastructure and knowledgeable MRI personnel challenges. A significant proportion of the high-field MRI scanners were in small-to-medium sized private centers, often unaffiliated with academic institutions and driven by a business model. Despite the general limited access to biomedical engineers to provide regular MRI equipment maintenance, a substantial proportion of low-field MRI scanners were still in clinical use past their obsolescence period. The prohibitive cost of MRI service contributes sigificantly to lack of access, significant underutilization, and challenges to sustainability of MRI in SSA. At an average cost of US$200 for an MRI examination and an average monthly salary of roughly US$470 purchasing power parity^39^, the average African in SSA cannot afford an MRI. In general, we discerned a zeal in the region to receive relevant training, research support, and access to collaborate and partner with the global MRI community to enable local experts and users realize the MRI needs in Africa, particularly Sub-Saharan Africa.

### State of MRI imaging in Sub-Saharan Africa

MRI in Africa has been evolving over the last decade, although the progress has been very slow and disproportionate in meeting the needs of the 1.5 billion people on the continent, and insufficient in competing adequately with global imaging trends. Africa, a continent with the largest global burden of chronic diseases and where premature deaths from non-communicable diseases, including heart disease and cancer, will surpass deaths from infectious diseases in 10 years^40^, has the worst access to MRI technology^27^. The USA with a population similar to West Africa has 37 times more MRI scanners than the entire region of West Africa^2,27^.

The CAMERA needs assessment survey (NAS) revealed that a considerable portion (40%) of MRI scanners in SSA are low field, that is <1 Tesla (T) field strength and 80% are still in use well past their obsolescence. These low-field units have limited resolution and poor signal-to-noise-ratio compared to 1.5T and 3T scanners standard in clinical practice and research but can be retooled to produce high-value images using machine learning^41^. Although the number of high-field scanners are steadily increasing in SSA (Figure 1), the severe shortage in skilled MRI personnel has exacerbated the lack of access to MRI and its advancement in the region to meet local needs.

MRI is grossly underutilized in SSA as our NAS indicated, with 47% of the sites performing clinical imaging on ∼5 patients/scanner/day and only 8% imaging 15 or more patients per/scanner/day. An average MRI facility in the Global North scans 20-23 patients/scanner/day and are able to do so because they have highly skilled MRI technologists, optimized MRI imaging protocols, access to MRI physicists and engineers, and highly specialized radiologists^42^. Around 65% of imaging facilities in SSA do not have access to an MRI physicist. This is quite troubling given that most available scanners provide low-quality images. There are roughly 2 radiologists per million people in SSA compared to 116 per million in Europe and North America^27^. These radiologists are non-subspecialized, work longer hours, face challenges of frequent machine breakdowns and low supply of consumables (e.g., contrast agents) and have to contend with sub-optimal infrastructure to care for a population with unique medical conditions^42^

For pre-clinical research, essential in drug discoveries, virtually none of these scanners have been used. In fact, a PubMed search revealed no publication from Africa where MRI was used for pre-clinical imaging. This is no surprise given that a large majority of pre-clinical research performed globally are on ≥ 3T MRI scanners. However, with novel imaging protocols and large animal models, African scientists supported by skills training can develop preclinical MRI research capacity on lower field strength scanners.

### Key challenges to advancement of MRI in biomedical sciences in Sub-Saharan Africa

The challenges in Africa are enormous and there is a critical need to build the necessary infrastructures, acquire adequate equipment, attract and retain qualified professionals, develop relevant educational training programs and have a political will that would generate policies for effective and accessible care within budgetary constraints. Given the growth of technological advances in imaging with the application of data science and artificial intelligence, without any robust intervention Africa’s disparities will worsen. The strategic solutions in Africa must be tailored to the unique socio-economic fabric, culture, and heterogeneous nature of the region. There is a huge temptation to dump ideas and innovations from developed nations on Africa. Even though this may appear simpler and faster, by and large the results over the years have been dismal at best. The solutions must come from within, through educational empowerment and capacity development.

Infrastructural problems such as lack of proper laboratories and unserviceable, obsolete equipment for research has hampered medical research in Africa, in general. This is even more acute for highly advanced and relatively expensive MRI scanners, which after 10 years are no longer deemed state-of-the art and receive no support from manufacturers and software developers^42^. For the growing fleet of newly acquired high-field scanners in SSA, this raises another area of concern as the majority will reach their end-of-life cycle by 2025. Lack of proper facilities cannot be understated as almost all scanners in SSA are not equipped with research licenses to unlock advanced research imaging sequences readily available in academic centers in other regions. Besides the principal challenge of skilled manpower, another key barrier to the advancement of MRI in SSA is the isolation of emerging local MRI experts. There are no formal networks within Africa to bring together African MRI experts and users to exchange ideas, pool resources, and develop relevant imaging solutions to address local needs. Although 72% of respondents to the CAMERA NAS participate in MRI research, an overwhelming majority are not able to engage or share their work with other MRI scientists nationally, regionally, or internationally (Figure 2). As of 2019, less than 1% of attendees to the ISMRM, the largest scientific society for development and application of MRI in biomedicine, come from Africa^43^. This is well reflected in MRI research output (publications) from Africa, which comprise less than 1% of global output. Moreover, major contributions from SSA come from either collaborative work with scientists from the “West” or institutions with a large presence of scientists from the “West” (e.g., The ENIGMA publications).

While the problems are vast and multidimensional, we aim to address 4 crucial challenges: access and availability, personnel training and education, research capacity and sustainable technology, using a four-prong approach within our framework summarized below in Figure 7.

### Anticipated Impact

MRI is a versatile imaging tool for investigating structural, functional and molecular changes in vivo. Improving training and research collaboration in SSA, will advance the use of MRI technology in the region to find relevant solutions to imaging needs, especially in the absence of ‘luxury suite’ imaging (e.g PET, MEG, high-density EEG)^3,27^. For instance, epilepsy disproportionately impacts SSA (10x higher incidence compared to high-income countries)^44^, and up to 30% of epilepsy patients require surgery to resect brain lesions to alleviate seizures. Anatomical brain MRI performed as standard of care to localize the lesion for surgery, appears normal in one-third of these patients and luxury suite imaging using PET or MEG and high-density EEG are functionally used to localize lesion(s)^45^. In SSA, these luxury technologies are extremely scarce; 3 PET scanners and no MEG in the entire region (minus South Africa)^27^. Functional MRI techniques (perfusion, diffusion spectroscopy) can functionally identify epileptic lesions^45^, but none of these are routinely used in clinics in the Global North and as such there are no standard image acquisition, processing, or interpretation guidelines Current research data reporting these techniques focus largely on 3T MRI studies using higher brain acquisition coils. By 1) training MRI radiographers to master acquisition of these advanced techniques and adapt parameters from research to their lower-field strength systems, 2) training MRI physicist to analyze images from their lower quality scanners leveraging on machine learning, 3) training biomedical engineers and physicists to build brain coils compatible with reconfigured and legacy systems to increase the spatial resolution, and 4) upskilling radiologists to read these novel MRI techniques, epilepsy surgical patients in SSA, may expect to have similar outcomes to their peers in the rest of the world. Until then, these African patients – ∼303,000/year^44^, will continue to suffer tractable epilepsy without any access to successful treatment. Similar remarks can be made for cancer, stroke, and cardiac imaging, as well research studies particularly in dementia and mental health where functional and molecular imaging biomarkers are highly sought after but underdeveloped in MRI scanners typically available in Sub-Saharan Africa.

## Conclusion

With nearly one-fifth of the world’s population, Africa has the least access to MRI machines. Our NAS combined with WHO and IAEA provide the most up-to-date data on MRI density in Africa, although several high-field systems have been installed since June 2020 when last survey responses were captured. Although the majority of responses came from West Africa, the findings provide unique insight into Africa’s MRI needs. While it appears that more high-field systems are being used in the region, reported gaps in training, maintenance, and research capacity indicate ongoing challenges in providing sustainable high value MRI access in SSA. As we lay out our roadmap for achieving access to high value MRI in SSA and establish task forces to effect our mission, we invite the global MRI community to partner with us, and our African colleagues in effecting transformative access to MRI.

## Data Availability

All data produced in the present study are available upon reasonable request to the authors

## Acknowledgements

The authors would like to thank the following individuals Uche Nwankwo, Okeji, and Kelechi Aguwanmba for assisting in distribution of the NAS through their radiographer associations and networks. The authors are grateful to all participants of the NAS study and to the moderators, speakers and attendees of the CAMERA symposia for providing valuable input. The authors thank Anu Gbadamosi, Head IT and Special Projects at Crestview Radiology Limited for providing images and overview of the clinic’s power and IT infrastructure. The authors would like to acknowledge the leadership of Professor Marion Smits, Past President of European Society of Magnetic Resonance in Medicine and Biology (ESMRMB) for supporting the establishment of CAMERA. We thank ESMRMB and ISMRM central offices for assistance in organizing the CAMERA symposia. Funding for this work and CAMERA is partly supported by the Chan Zuckerberg Initiative (CZI) Expanding Global Access to Bioimaging RFA (2021-240505, UCA, GO, JO, ECN) and the Healthy Brain and Healthy Lives (HBHL) New Recruit Start-Up Supplements (2b-NISU-17, UCA).

## Author Contributions

Udunna C Anazodo conceptualized and developed the outline of the manuscript. Udunna C Anazodo, Jinggang J Ng, Boaz Ehiogu, Iris Asllani, and Farouk Dako conducted background work, analyzed the NAS and wrote the first draft. Udunna C Anazodo designed the NAS along with editorial contributions from the CAMERA Environmental Scan Task Force (Edward Chege Nganga, Godwin Ogbole, Farouk Dako, Henk-Jan Mustaerts, Mario Forjaz Secca, and Johnes Obungoloch). All authors provided valuable input, reviewed, and approved the final version of the manuscript.

## References

1. Geethanath S, Vaughan JT. Accessible magnetic resonance imaging: A review. J Magn Reson Imaging. 2019;49(7):e65–e77. doi:10.1002/jmri.26638

2. Ogbole GI, Adeyomoye AO, Badu-Peprah A, Mensah Y, Nzeh DA. Survey of magnetic resonance imaging availability in West Africa. Pan Afr Med J. 2018;30:240. doi:10.11604/pamj.2018.30.240.14000

3. Hricak H, Abdel-Wahab M, Atun R, et al. Medical imaging and nuclear medicine: a Lancet Oncology Commission. Lancet Oncol. 2021;22(4):e136–e172. doi:10.1016/S1470-2045(20)30751-8

4. Frumkin H, Haines A. Global Environmental Change and Noncommunicable Disease Risks. Annu Rev Public Health. 2019;40:261–282. doi:10.1146/annurev-publhealth-040218-043706

5. Milanzi E, Nkoka O, Kanje V, Ntenda P. Air pollution and non-communicable diseases in Sub-Saharan Africa. Sci African. 2021;11:e00702. doi:10.1016/j.sciaf.2021.e00702

6. Ajayi IO, Adebamowo C, Adami H-O, et al. Urban-rural and geographic differences in overweight and obesity in four sub-Saharan African adult populations: a multi-country cross-sectional study. BMC Public Health. 2016;16(1):1126. doi:10.1186/s12889-016-3789-z

7. Gyasi RM, Phillips DR. Aging and the Rising Burden of Noncommunicable Diseases in Sub-Saharan Africa and other Low-and Middle-Income Countries: A Call for Holistic Action. Gerontologist. 2020;60(5):806–811. doi:10.1093/geront/gnz102

8. Gouda HN, Charlson F, Sorsdahl K, et al. Burden of non-communicable diseases in sub-Saharan Africa, 1990&–2017: results from the Global Burden of Disease Study 2017. Lancet Glob Heal. 2019;7(10):e1375–e1387. doi:10.1016/S2214-109X(19)30374-2

9. Kroll C, Warchold A, Pradhan P. Sustainable Development Goals (SDGs): Are we successful in turning trade-offs into synergies? Palgrave Commun. 2019;5(1):140. doi:10.1057/s41599-019-0335-5

10. Mensah GA, Roth GA, Fuster V. The Global Burden of Cardiovascular Diseases and Risk Factors: 2020 and Beyond. J Am Coll Cardiol. 2019;74(20):2529–2532. doi:10.1016/j.jacc.2019.10.009

11. Sliwa K, Ntusi N. Battling Cardiovascular Diseases in a Perfect Storm. Circulation. 2019;139(14):1658–1660. doi:10.1161/CIRCULATIONAHA.118.038001

12. Ntusi N. Cardiovascular disease in sub-Saharan Africa. SA Hear. 2021;18(2):74–77. doi:10.10520/ejc-saheart-v18-n2-a1

13. Borghammer P, Jonsdottir KY, Cumming P, et al. Normalization in PET group comparison studies--the importance of a valid reference region. Neuroimage. 2008;40(2):529–540. doi:10.1016/j.neuroimage.2007.12.057

14. Qin C, Murali S, Lee E, et al. Sustainable low-field cardiovascular magnetic resonance in changing healthcare systems. Eur Heart J. 2022:1–15. https://doi.org/10.1093/ehjci/jeab286.

15. Bernasconi A, Cendes F, Theodore WH, et al. Recommendations for the use of structural magnetic resonance imaging in the care of patients with epilepsy: A consensus report from the International League Against Epilepsy Neuroimaging Task Force. Epilepsia. 2019;60(6):1054–1068. doi:10.1111/epi.15612

16. Bhat SS, Fernandes TT, Poojar P, et al. Low-Field MRI of Stroke : Challenges and Opportunities. J Magn Reson Imaging. 2020. doi:10.1002/jmri.27324

17. Okorie CK, Ogbole GI, Owolabi MO, Ogun O, Adeyinka A, Ogunniyi A. Role of Diffusion-weighted Imaging in Acute Stroke Management using Low-field Magnetic Resonance Imaging in Resource-limited Settings. West African J Radiol. 2015;22(2):61–66. doi:10.4103/1115-3474.162168

18. Frisoni GB, Fox NC, Jack CRJ, Scheltens P, Thompson PM. The clinical use of structural MRI in Alzheimer disease. Nat Rev Neurol. 2010;6(2):67–77. doi:10.1038/nrneurol.2009.215

19. Akinyemi RO, Okubadejo N, Yaria J, et al. Dementia in Africa : Current evidence, knowledge gaps, and future directions. Alzheimer’s Dement. 2021;(February):1–20. doi:10.1002/alz.12432

20. Justice O, Jordan LC, Lee CA, et al. Safety of 3 Tesla Magnetic Resonance Imaging in Patients with Sickle Cell Disease. COSTA ALF, ed. Radiol Res Pract. 2021;2021:5531775. doi:10.1155/2021/5531775

21. Maude RJ, Barkhof F, Hassan MU, et al. Magnetic resonance imaging of the brain in adults with severe falciparum malaria. Malar J. 2014;13(1):177. doi:10.1186/1475-2875-13-177

22. Mallon D, Doig D, Dixon L, Gontsarova A, Jan W, Tona F. Neuroimaging in Sickle Cell Disease: A Review. J Neuroimaging. 2020;30(6):725–735. doi:https://doi.org/10.1111/jon.12766

23. Brown LC, Ahmed HU, Faria R, et al. Multiparametric MRI to improve detection of prostate cancer compared with transrectal ultrasound-guided prostate biopsy alone: the PROMIS study. Health Technol Assess. 2018;22(39):1–176. doi:10.3310/hta22390

24. Gatta G, Trama A, Capocaccia R. Variations in cancer survival and patterns of care across Europe: roles of wealth and health-care organization. J Natl Cancer Inst Monogr. 2013;2013(46):79–87. doi:10.1093/jncimonographs/lgt004

25. RAD-AID. Radiology-Readiness: A Framework for Implementing Radiology in Resource-Limited Regions. https://rad-aid.org/resource-center/radiology-readiness. Accessed February 21, 2020.

26. Global Health Observatory data repository. World Health Organization. https://apps.who.int/gho/data/node.main.510. Published 2016. Accessed August 4, 2020.

27. Human Health Campus. IMAGINE - IAEA Medical imAGIng and Nuclear mEdicine global resources database. International Atomic Energy Agency (IAEA). https://humanhealth.iaea.org/HHW/DBStatistics/IMAGINE.html. Accessed August 4, 2020.

28. Journal Citation Reports. 2020 Journal Impact Factor. 2021.

29. Ntshebe O. Sub-Saharan Africa’s Brain Drain of Medical Doctors to the United States: An Exploratory Study. Insight on Africa. 2010;2(2):103–111. doi:10.1177/0975087814411123

30. Raji A, Joel A, Ebenezer J, Attah E. The Effect of Brain Drain on the Economic Development of Developing Countries: Evidence from Selected African Countries. September 2018.

31. Emetere ME, Agubo O, Chikwendu L. Erratic electric power challenges in Africa and the way forward via the adoption of human biogas resources. Energy Explor Exploit. 2021;39(4):1349–1377. doi:10.1177/01445987211003678

32. Elahi A, Dako F, Zember J, et al. Overcoming Challenges for Successful PACS Installation in Low-Resource Regions: Our Experience in Nigeria. J Digit Imaging. 2020;33(4):996–1001. doi:10.1007/s10278-020-00352-y

33. Harper JR, Cherukuri V, O’Reilly T, et al. Assessing the utility of low resolution brain imaging: treatment of infant hydrocephalus. NeuroImage Clin. 2021;32:102896. doi:https://doi.org/10.1016/j.nicl.2021.102896

34. Marques JP, Simonis FFJ, Webb AG. Low-field MRI: An MR physics perspective. J Magn Reson Imaging. 2019;49(6):1528–1542. doi:10.1002/jmri.26637

35. Alexander DC, Zikic D, Ghosh A, et al. Image quality transfer and applications in diffusion MRI. Neuroimage. 2017;152:283–298. doi:https://doi.org/10.1016/j.neuroimage.2017.02.089

36. Lin H, Figini M, Tanno R, et al. Deep Learning for Low-Field to High-Field MR: Image Quality Transfer with Probabilistic Decimation Simulator. Lect Notes Comput Sci. 2019;11905 LNCS:58-70. doi:10.1007/978-3-030-33843-5_6

37. Mollura DJ, Culp MP, Pollack E, et al. Artificial Intelligence in Low-and Middle-Income Countries: Innovating Global Health Radiology. Radiology. 2020;297(3):513–520. doi:10.1148/radiol.2020201434

38. GE Healthcare. Kenyan Small and Medium Healthcare providers to get boost in accessing financing for Medical Equipment purchases. https://ge.africa-newsroom.com/press/kenyan-small-and-medium-healthcare-providers-to-get-boost-in-accessing-financing-for-medical-equipment-purchases. Published 2019. Accessed February 2, 2020.

39. Stanwix B. What do minimum wages look like in sub-Saharan Africa? World Econ Forum. 2015. https://www.weforum.org/agenda/2015/10/what-do-minimum-wages-look-like-in-sub-saharan-africa/. Accessed March 2, 2022.

40. Ezzati M, Pearson-Stuttard J, Bennett JE, Mathers CD. Acting on non-communicable diseases in low-and middle-income tropical countries. Nature. 2018;559(7715):507–516. doi:10.1038/s41586-018-0306-9

41. Figini M, Lin H, Ogbole G, et al. Image Quality Transfer Enhances Contrast and Resolution of Low-Field Brain MRI in African Paediatric Epilepsy Patients. arXiv Prepr 200307216. 2020.

42. Zhang L, Hefke A, Figiel J, Schwarz U, Rominger M, Klose KJ. Enhancing Same-Day Access to Magnetic Resonance Imaging. J Am Coll Radiol. 2011;8(9):649–656. doi:10.1016/j.jacr.2011.04.001

43. Warnert EAH, Kasper L, Meltzer CC, et al. Resonate: Reaching Excellence Through Equity, Diversity, and Inclusion in ISMRM. J Magn Reson Imaging. 2021;53(5):1608–1611. doi:10.1002/jmri.27476

44. Kissani N, Nafia S, El Khiat A, et al. Epilepsy surgery in Africa: state of the art and challenges. Epilepsy Behav. 2021;118:107910. doi:10.1016/j.yebeh.2021.107910

45. Poirier SE, Kwan BYM, Jurkiewicz MT, et al. 18F-FDG PET-guided diffusion tractography reveals white matter abnormalities around the epileptic focus in medically refractory epilepsy : implications for epilepsy surgical evaluation. Eur J Hybrid Imaging. 2020;4(10). doi:10.1186/s41824-020-00079-7

