## Supplementary Materials for "A Framework for Advancing Sustainable MRI Access in Africa"

**Table S1: Appended CAMERA NAS Survey**

The Survey is appended at the end of this supplementary material.

**Table S2: MRI Scanner Density aggregated from CAMERA NAS, IAEA IMAGINE and WHO Medical Device datasets**

| Country | MRI Density (pmp) | Country Cont'd | MRI Density (pmp) |
|---|---|---|---|
| Algeria | 3.484 | Liberia | 0.000 |
| Angola | 0.031 | Libya | 4.869 |
| Benin | 0.085 | Madagascar | 0.000 |
| Botswana | 0.434 | Malawi | 0.107 |
| Burkina Faso | 0.049 | Mali | 0.000 |
| Burundi | 0.000 | Mauritania | 0.884 |
| Cape Verde | 1.818 | Mauritius | 4.740 |
| Cameroon | 0.039 | Morocco | 3.290 |
| CAR | 0.000 | Mozambique | 0.033 |
| Chad | 0.000 | Namibia | 0.802 |
| Congo | 0.372 | Niger | 0.000 |
| Cote d'Ivoire | 0.117 | Nigeria | 0.303 |
| Djibouti | 1.027 | Rwanda | 0.079 |
| DR Congo | 0.000 | Senegal | 0.123 |
| Egypt | 1.942 | Seychelles | 10.243 |
| Eritrea | 0.286 | Somalia | 0.130 |
| Ethiopia | 0.062 | Sierra Leone | 0.128 |
| Gabon | 0.921 | South Africa | 2.630 |
| Gambia | 0.426 | Sudan | 0.000 |
| Ghana | 0.526 | Tanzania | 0.000 |
| Guinea | 0.737 | Togo | 0.124 |
| Guinea-Bissau | 0.000 | Tunisia | 4.275 |
| Kenya | 0.951 | Uganda | 0.068 |
| Lesotho | 0.471 | Zambia | 0.056 |
| *pmp = per million people* | | Zimbabwe | 0.273 |

**Table S3: Personnel Distribution in MRI Centers from CAMERA NAS**

| Total Number | Radiologist | Radiographer | Physicist |
|---|---|---|---|
| None | 1 | 4 | 57 |
| 1 Staff | 10 | 7 | 14 |
| 2-5 Staff | 46 | 47 | 13 |
| 6-10 Staff | 16 | 19 | 3 |
| >10 Staff | 14 | 10 | 0 |
| Cumulative Total | 87 | 87 | 87 |

**Table S4: Research Capacity based on Total number of Publications**

| | Total Number of Countries | Total Publication Count | % of Total over 3-Year |
|---|---|---|---|
| All LMIC | 14 | 104 | 1.28 |
| All UMIC | 22 | 930 | 11.44 |
| All HIC | 47 | 7097 | 87.28 |
| Africa | 11 | 40 | 0.49 |
| SSA | 7 | 17 | 0.21 |

*LMIC = Low-middle income countries; UMIC = Upper middle-income countries; HIC = High income countries; SSA = Sub-Saharan Africa. Data was summarized from the Journal of Citation Reports Journal Profile based on research contributions by country/region in the most three-year period from 2020 (2020 Journal Impact Factor, Journal Citation Reports (Clarivate, 2021)). An excel sheet of the full list of research contributions by country is appended in supplementary materials 2.*

**Table S5: Strategy to increase MRI access and research translation in Africa.**

| Challenge Theme/Target | Challenges | Opportunities/Solutions |
|---|---|---|
| Access and Availability | • High equipment costs<br>• Limited access to financial capital for equipment procurement and maintenance<br>• Poor reimbursement and predominant out of pocket payment option<br>• Limited access to stable electricity and power | • Special vendor pricing for Africa<br>• Vendor financing programs<br>• Increase public and private investment in medical device manufacturing<br>• Healthcare-specific financing funds |

| | | |
|---|---|---|
| | infrastructure for high-field MRI<br>● Limited access to image post-processing and picture archiving and communication system (PACS) for rapid diagnosis and image analysis<br>● Limited access to Internet, network connectivity and other related technology for MRI data management | ● Increase public medical insurance coverage and improve utilization of healthcare insurance<br>● Investment in low-field MRI<br>● Utilization of AI technology to enhance low-field MRI, provide high value MRI service, and extend life cycle of high field MRI<br>● Sourcing locally-produced medical equipment and consumables to lower costs<br>● Utilization of compact and temperature resistant portable servers to eliminate need for expensive climate-controlled server rooms<br>● Utilization of MiniPACs to enable local data storage when cloud connectivity is limited or unavailable<br>● Schedule cloud backup for non-peak hours when bandwidth usage is low to reduce network demands |
| Personnel Training | ● Scarcity of trained technicians, radiologists, and biomedical engineers to run, effectively utilize, and maintain MRI scanners<br>● Lackluster medical manufacturing market perpetuates poor technical expertise among local populations<br>● Africa’s brain drain reduces availability of medical professionals in the region | ● Intervention of ISMRM, ESMRMB, CAMERA and OEMs to set up curriculum-based training programs, mentorship and education opportunities<br>● Improve OEM and MRI users/personnel engagement<br>● Investment in low-field MRI reduces technical demands |

| | | |
|---|---|---|
| | | • Creation of inter-regional network and global partnerships to support and retain MRI experts in the region<br>• Integration of AI education with conventional radiology training to enable the safe utilization of AI technology by local radiologists |
| Research Capacity | • Low production of medical research resulting in poor knowledge surrounding best clinical practices in low resource settings<br>• High proportion of private to academic MRI facilities who have limited research capacity<br>• Lack of data diversity in medical imaging research from Africa<br>• Limited access to electronic medical records | • Increase public and private investment in medical research<br>• Create regional research hubs to link private centers to local/regional academic institutions<br>• Create inter-regional network and global partnerships to link MRI experts in the region<br>• Digitization of radiology to enhance access to medical data<br>• Adoption of enterprise imaging systems to enable inclusion of data from private clinics<br>• Development of regulatory systems for health data management |
| Sustainable Technology | • Lack of regulatory policy and facility quality standards<br>• Lack of technical expertise to maintain MRI equipment<br>• Unclean electricity may damage expensive equipment | • Development of legal and regulatory frameworks to enhance procurement, establish facility quality standards and support medical research<br>• Develop local technical expertise to maintain and sustain MRI equipment and effectively extend life cycle<br>• Coupling up of energy sources to provide reliable electricity |

**Table S6: Short Answer Responses to Optional Questions of the CAMERA NAS**

A) Challenges faced by your facility that impair day to day operation of the MRI program

1. *Unavailability of consumables*
2. *Power fluctuations, software problems, problems with cooling system*
3. *Delays in gas replacement*
4. *Power supply problems; repairs take long because engineer stays in another country; service contracts become too expensive as machine gets older resulting into spare parts becoming too expensive*
5. *There is generalized wide knowledge gap among referring doctors making it difficulty to achieve effective MRI scan and result for proper diagnosis and patient management which most often result in repeat scans. Also the radiologists are not skilled in MRI film reporting. The request form are poorly filled and most times are blank. Patient waiting time is unduly long resulting in patient dissatisfaction. Arbitrary price of MRI procedures due to very few( only 2) functioning machines servicing 6 states of about 20million population etc. Organizational beuracracy that pays more interest to revenue than quality etc.*
6. *Power supply, lack of spare parts*
7. *Awareness of the capacity and capabilities of the facility. Patient turn out is low, possibly due to it's location and awareness as well as cost*
8. *We do not have MRI compatible Anaesthetic monitoring devices thereby unable to attend to paediatric and non-co-operating patient.*
9. *Application softwares difficult to come by.Maintenance culture quite insufficient.Problem of steady power supply.Lack of standardised PACS facility*
10. *Not all pulse sequences are activated and we work with limited coils due to the challenge of the overall cost.*

B. Challenges that impair facilities ability to conduct or participate in MRI-related research

11. *Busy working schedules of staff*
12. *The lack of funds and technical know-how*
13. *Limited coils and pulse sequences*
14. *They are business-minded*
15. *Poor record keeping due to manual methods applied.*
16. *Proper acquisition of image that can be compared with the ones in the western world*

17. *Incomplete information, no back up system*

18. *No Research Board, no sponsorship or grant facilities, too much emphasis on returns on investment.*

19. *Using a single scanner. It makes it difficult to go into research when you have patients that really need the machine.*

20. *We dont have and cannot employ people for research*

C. Challenges in personnel capacity that impair ability to perform MRI well

21. *In my point of view, the quality of our exam is good, but we use 0.4 Tesla and it is not performed to conducted all the parts of the body l'île breast, abdomen, thoracic MRI.*

22. *High attrition rate of radiographers*

23. *Lack of training, frequent equipment break down*

24. *Lack of training is the biggest problem. No institution provides MRI training locally. No MRI short courses available within easy reach*

25. *Engineers that repair the machines are inadequate*

26. *Training and experience. The low patient inflow as not given us enough experience and exposure.*

27. *At the moment, with the newly installed MRI and experienced staff, we are good.*

28. *No in-house engineers for regular servicing.*

29. *Relatively poor competence in rare MRI studies*

30. *We have only two personnel who have achieved some level of training. This is also the first government own MRI in the country. It would be good if more people could be trained.*

D. Solution for tackling challenges faced in using MRI clinically

31. *Employ and train more radiographers and motivate them well*

32. *Good power supply and continuous clinical education*

33. *Training of staffs and the need for planned preventive maintenance*

34. *The lack of refresher course on MRI physics short courses available within easy reach*
35. *Availability of an MRI training Institute / Availability of MRI (online) update courses/ Creation*

*of MRI technologists association where techs meet and discuss challenges and possible solutions*

36. *Training and exposure. / Our centre is a young centre with no much experience.*
37. *Provision of more than one scanner in the unit*
38. *Have more than two scanners and effective mode of power supply.*
39. *Purchase of MRI scanner with at least 3Tesla. Sponsorship of Radiologists and radiographers for conferences and CME*
40. *Provision of Higher Tesla Scanner / Good Electricity supply / Improved Training programs for personnel*

E. Solution for tackling challenges faced in conducting MRI research

41. *Collect cases of studies day after day.*
42. *Availability of funds and access to research information*
43. *Educating the team about the importance of research in the field of mri.*
44. *Appropriate systematic medical record keeping, PACS and RIS to aid clinical research.*
45. *Training programs and spending time at centres with high volume turn out and involving in research programs with others in order to learn and be motivated*
46. *GETTING THE COMMITTED AND RIGHT PARTNERSHIP.*
47. *Collaboration with centres with better magnets*
48. *Provision of higher Tesla scanner / Motivation for staff in terms of research grants*
49. *Train more MRI radiographers. Extend working hours. Pay radiographers for research or make them co-authors*
50. *Provision of coils and payment for the activation of other pulse sequences by the government or philanthropist.*

*Unedited randomly selected responses from 70 NAS respondents.*

**Figure S1: Reported MRI Research Topic Interests from the CAMERA NAS**

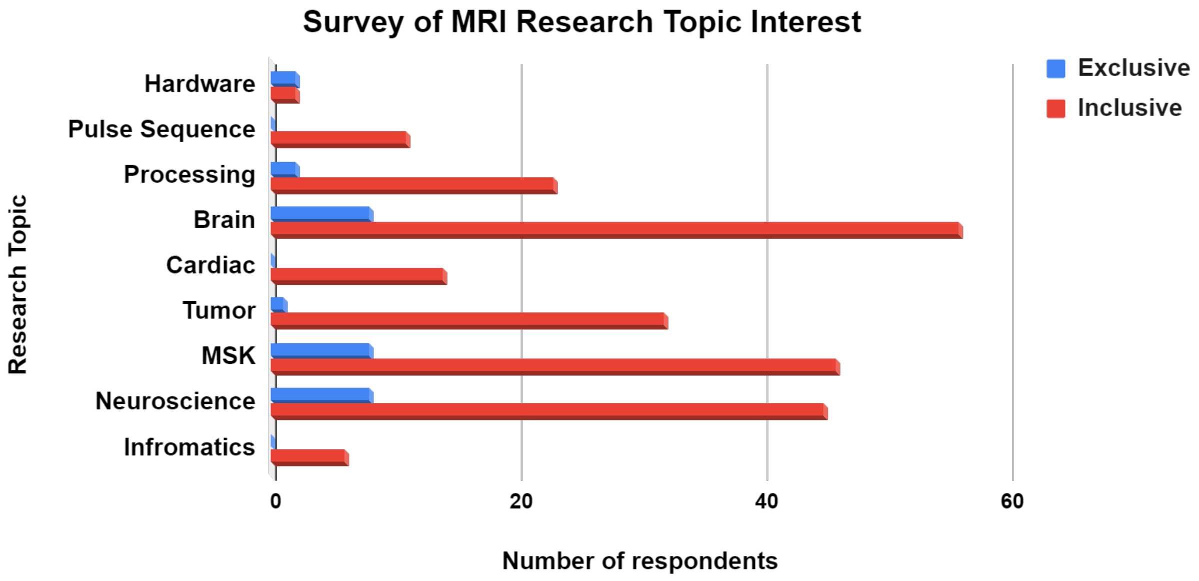


*A total of 87 respondents indicated exclusive (only topic selected) or inclusive interest in MRI physics hardware (scanner/coil development), MRI physics sequence development, image processing, brain imaging application, cardiac imaging application, tumor imaging application, musculoskeletal (MSK) imaging application, neuroscience or informatics/radiomics.*

**Figure S2: Research Output in the Global Context**

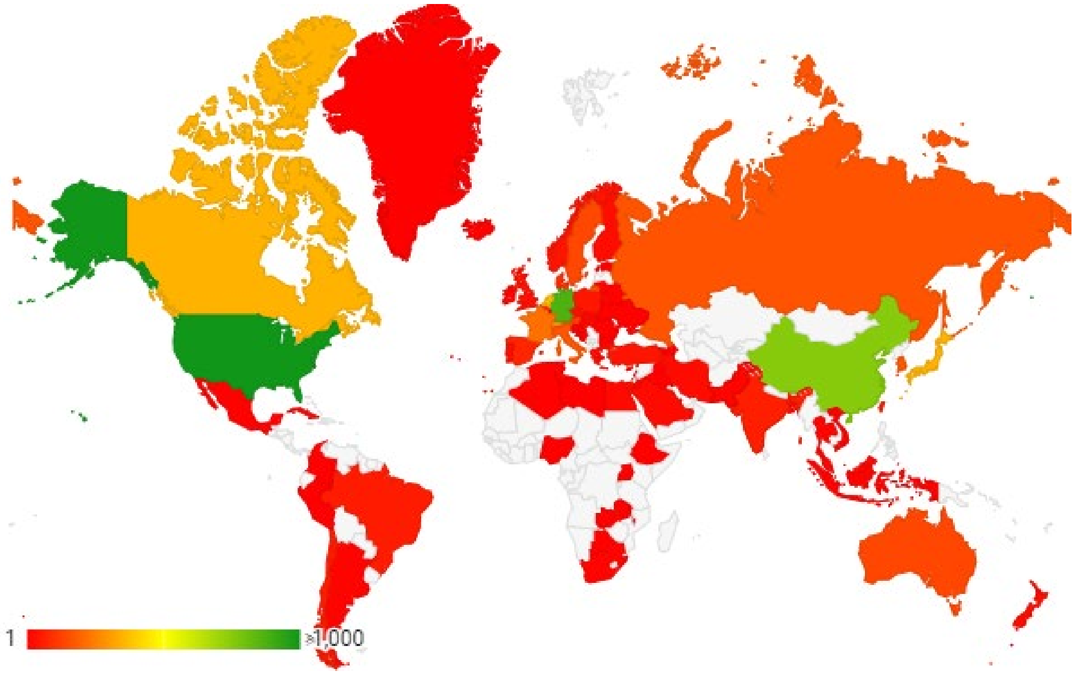


*Total number of MRI studies published in MRI exclusive journals between 2018 and 2020 for each country with data (colored).*

**Figure S3: Most Common Reported Clinical Indications for MRI**

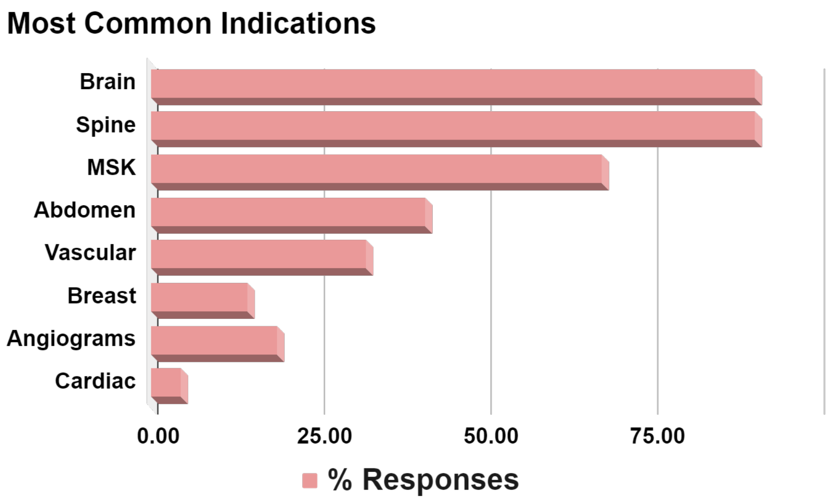


*Percent of a total 87 CAMERA NAS responders.*

**Figure S4: Funding Sources for Payment of MRI Services**

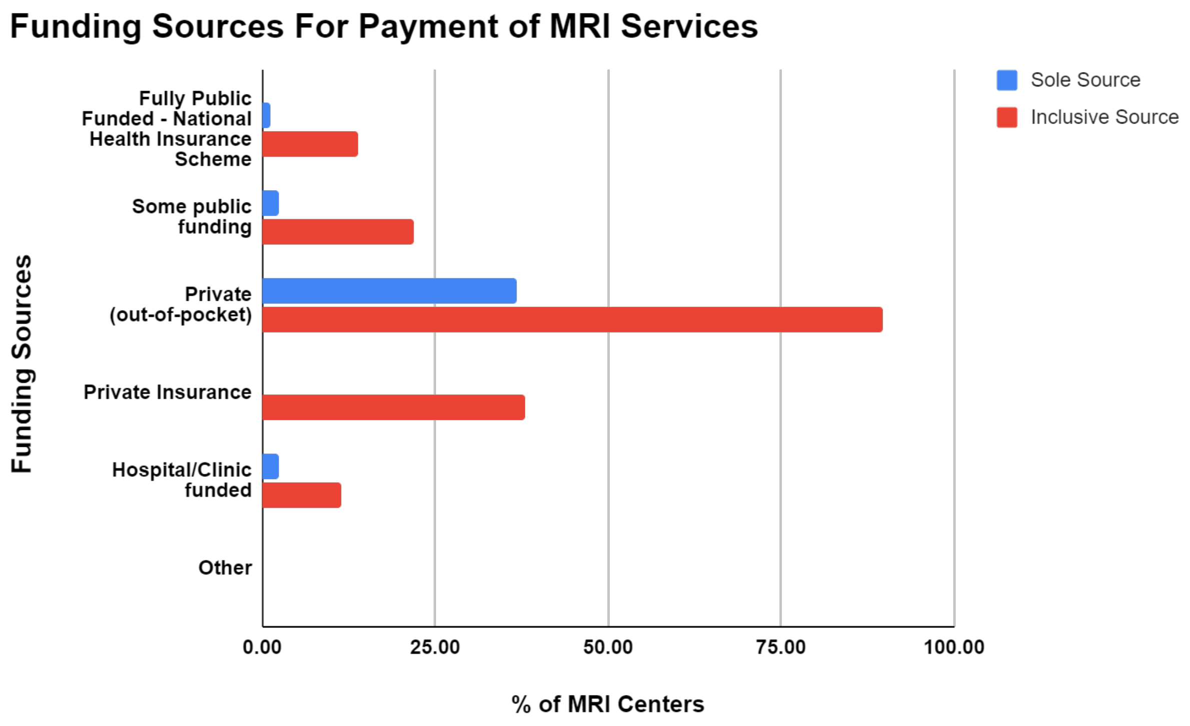


*The breakdown of sources of funding to pay for MRI services as a percent of the 87 MRI centers reporting data. Responses are shown here indicating responders that reported one of the payment options (sole source) and responders that reported two or more sources of payment (inclusive source).*

#### Figure S5: Energy Solutions for Steady Power Supply in SSA Medical Imaging Facilities

An illustration of the energy solutions at Crestview Radiology Clinic, Lagos, Nigeria for steady power supply to support operation of an MRI and other imaging devices.

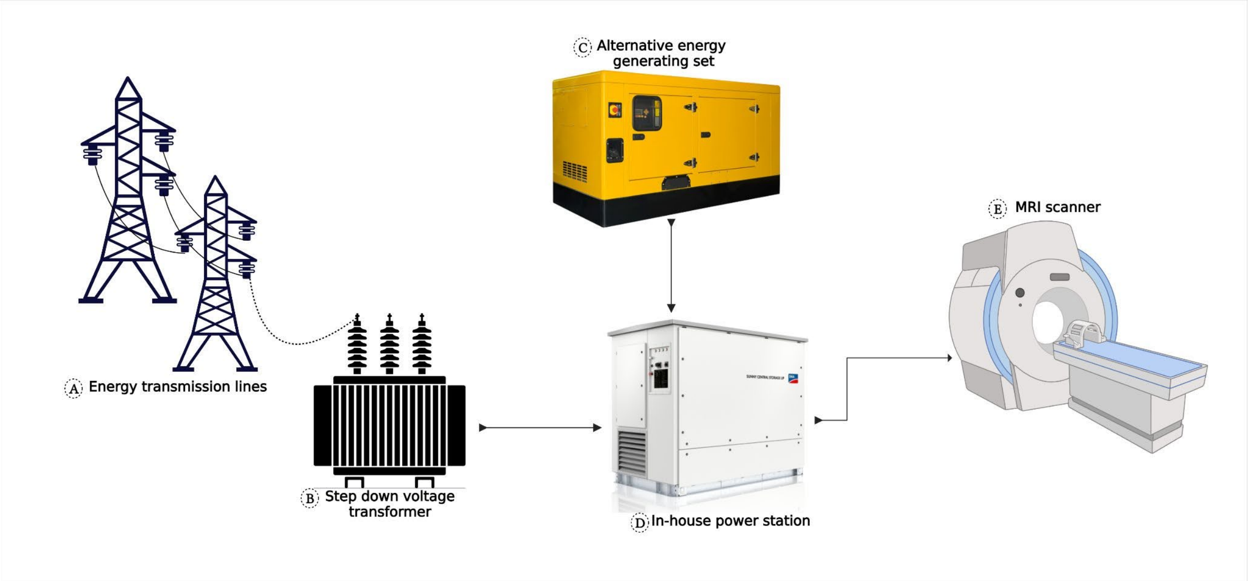


#### Figure S6: CAMERA Governance Structure

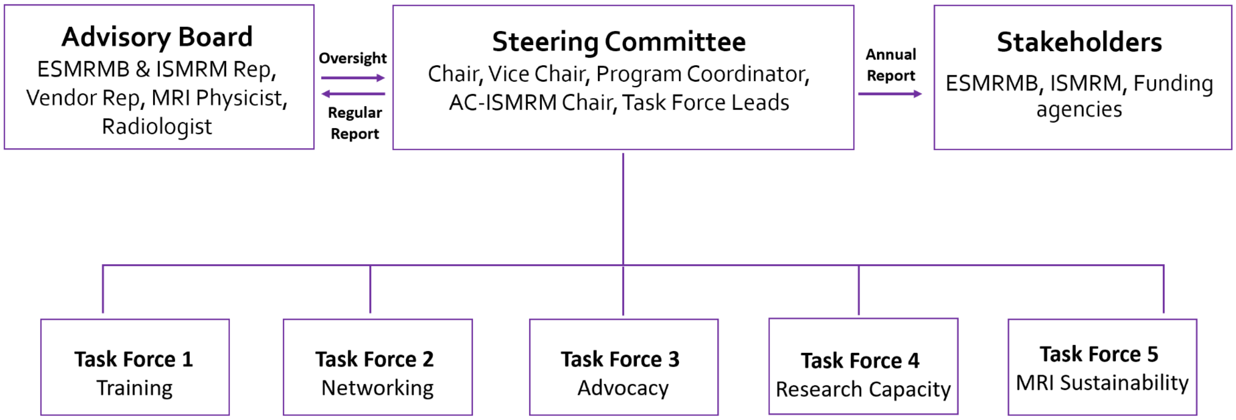

### MRI Scanner and Personnel Survey

Dear Radiologist or Radiographer or Physicist or Scientist,

Thank you in advance for taking time to complete this survey from the Committee for Advancement of MRI Education and Research in Africa (CAMERA). CAMERA is a Working Group of the European Society for Magnetic Resonance in Medicine and Biology (ESMRMB) that is working to develop a sustainable framework for accelerating MRI research and education excellence in Africa, a region of the world with the lowest density of MRI scanners.

This 15 minute survey is the first step towards achieving CAMERA's vision of reducing the disparity in MRI research capacity in Africa.

By completing this survey, you will help CAMERA:
1. Identify and understand needs and challenges of conducting MRI for clinical use and research in Africa.

2. Identify and understand gaps in training of highly qualified personnel in MRI research and clinical practice.

3. Create educational programs to help close gaps in training of highly qualified personnel in MRI.

4. Help create a consortium of MRI experts and users from Africa interested in collaborating with African MRI scientists and global partners.

5. Help our vendor partners know how to support MRI needs in Africa.

All information provided in this survey is confidential and accessible solely to the members of the CAMERA MRI listed at the end of this survey.

If you have questions about this survey, please contact:.

* Required

1. Email *

______________________________

#### Facility Information

2. Please indicate your specialty or designation *

*Mark only one oval.*

- Radiologist
- Radiographer / Technologist
- Physicist
- Other: ______________________

3. Where is your Diagnostic Imaging facility located? Please select only one. *

*Mark only one oval.*

- Algeria
- Angola
- Benin
- Bostwana
- Burkina Faso
- Burundi
- Cameroon
- Canary Islands
- Cape Verde
- Central African Republic
- Chad
- Cote d'Ivoire
- Democratic Republic of the Congo
- Djibouti
- Egypt
- Equatorial Guinea
- Eritrea
- Ethiopia
- Gabon
- Gambia
- Ghana
- Guinea
- Guinea-Bissau
- Kenya
- Lesotho
- Liberia
- Libya
- Madagascar
- Malawi
- Mali

- [illegible]
- Morocco
- Mozambique
- Namibia
- Niger
- Nigeria
- Republic of Congo
- Rwanda
- Senegal
- Sierra Leone
- Somalia
- South Africa
- Sudan
- Tanzania
- Togo
- Tunisia
- Uganda
- Zambia
- Zimbabwe

4. Please describe your facility ownership and affiliations. Please select all that apply. *

*Check all that apply.*

- [ ] Public/Government
- [ ] Private practice/out-patient only radiology
- [ ] Non-Profit including religious charities
- [ ] University Hospital
- [ ] Community hospital or health care center or ambulatory clinic
- [ ] Tertiary or Regional Hospital/ Health Care Center

Other: [ ] ____________________

5. Please indicate the number of personnel you have in your facility *

*Mark only one oval per row.*

| | None | 1 | 2-5 | 6-10 | >10 |
|---|---|---|---|---|---|
| Radiologists | ◯ | ◯ | ◯ | ◯ | ◯ |
| MRI Radiographer / technologists | ◯ | ◯ | ◯ | ◯ | ◯ |
| MRI Physicists | ◯ | ◯ | ◯ | ◯ | ◯ |
| Nursing Staff | ◯ | ◯ | ◯ | ◯ | ◯ |
| Other Staff | ◯ | ◯ | ◯ | ◯ | ◯ |

6. Do you have an MRI Scanner at your Imaging Facility or Department? *

*Mark only one oval.*

◯ Yes

◯ No *Skip to question 38*

#### Scanner Description

7. What is the total number of MRI scanners at your at your center *

_______________________________

8. How many of the total number of MRI scanners are working (not broken) and in use at the moment *

*Mark only one oval.*

- 0
- 1
- 2
- 3
- 4
- 5
- All

9. Select the total number of scanners for each scanner type you have at your center. Select ONLY what applies to your facility

*Check all that apply.*

| | 0 | 1 | 2 | 3 | 4 | 5 |
|---|---|---|---|---|---|---|
| 3 Tesla | ☐ | ☐ | ☐ | ☐ | ☐ | ☐ |
| 1.5 Tesla | ☐ | ☐ | ☐ | ☐ | ☐ | ☐ |
| 1.0 Tesla | ☐ | ☐ | ☐ | ☐ | ☐ | ☐ |
| <1.0 Tesla | ☐ | ☐ | ☐ | ☐ | ☐ | ☐ |

10. Describe your scanner, make and field strength. Select ONLY what applies to your facility

*Check all that apply.*

| | Toshiba/Canon | Siemens | General Electric | Philips | Other |
|---|---|---|---|---|---|
| 3 Tesla | ☐ | ☐ | ☐ | ☐ | ☐ |
| 1.5 Tesla | ☐ | ☐ | ☐ | ☐ | ☐ |
| 1.0 Tesla | ☐ | ☐ | ☐ | ☐ | ☐ |
| <1.0 Tesla | ☐ | ☐ | ☐ | ☐ | ☐ |

11. How long have you had your scanner? Select ONLY what applies to your facility

*Check all that apply.*

| | 5-10 years | 3-4 years | 1-2 years | < 1 Year |
|---|---|---|---|---|
| 3 Tesla | ☐ | ☐ | ☐ | ☐ |
| 1.5 Tesla | ☐ | ☐ | ☐ | ☐ |
| 1.0 Tesla | ☐ | ☐ | ☐ | ☐ |
| <1.0 Tesla | ☐ | ☐ | ☐ | ☐ |

12. Does your facility have access to a Service Engineer who takes care of technical issues with the MRI scanner *

*Mark only one oval.*

( ) Yes

( ) No *Skip to question 15*

#### Maintenance

13. If you have a service engineer, is the engineer *

*Check all that apply.*

- [ ] A staff of the facility/insitution/clinic
- [ ] from a 3rd party service provider
- [ ] Other

14. If yes, how often does your Service Engineer perform scheduled and regular performance maintenance of your scanner *

*Check all that apply.*

- [ ] Monthly
- [ ] up to 2-3 times per year
- [ ] Once per year - Annually
- [ ] Never - only when the scanner is broken or down

#### Usage

15. What is the approximate total number of patients that you scan on your MRI at your facility per day *

*Mark only one oval.*

- ( ) up to 5
- ( ) 6-10
- ( ) 10-15
- ( ) 15+

16. Downtime Frequency. How often is your MRI scanner unavailable for use or down *

*Mark only one oval.*

- Often (1-2 times per week)
- Occasional (1-3 times per month)
- Seldom (<1 time in a 6 month span)
- Never

17. What are the most common indications for MRI scans at your facility? Choose all that apply. *

*Check all that apply.*

- [ ] Brain - Routine, Seziure, Orbits, Headaches/Migraine, Space Occupying lesions
- [ ] Angiograms
- [ ] Cardiac - Ischaemia, infiltrative disease, congential malformation, cardiomayopathy
- [ ] Musculoskeletal (MSK) - Wrist, Shoulder, Knee, Ankle, Hip, Long bones
- [ ] Breast
- [ ] Cerebrovascular and Vascular studies
- [ ] Abdomen and Pelvis - Liver, Kidney, Basic Abdomen
- [ ] Spine - Cervical, Thoaraic and Lumbo-Sacral

#### Facility Infrastructure

18. How often is the power supply at your facility? Is power available... *

*Check all that apply.*

- [ ] 100% of the time
- [ ] 75% (3/4) of the time
- [ ] 50% (1/2 to 3/4) of the time
- [ ] Less than 50% of the time

19. Do you have a source of back- up power

*Mark only one oval.*

- Yes
- No

20. Does your facility have a Picture Archiving and Communication System (PACS) to store and retrieve images

*Mark only one oval.*

- Yes
- No

21. Does your facility use teleradiology? Teleradiology is the interpretation and/or consultation of MRI acquired at your facility at another location

*Mark only one oval.*

- Yes
- No

#### Personnel Training and Education

22. If you have a Radiologist at your Facility, how often does the Radiologists at your facility engage in continuing medical education each year

*Mark only one oval.*

- I don't know
- Often (at least 2-3 times per year)
- Seldom (once per year)
- Never

23. How often does the Radiographer/Technologists at your facility engage in continuing education each year

*Mark only one oval.*

- I don't know
- Often (at least 2-3 times per year)
- Seldom (once per year)
- Never

24. If you have a Radiologist at your Facility, how often does your Radiologists attend the following forms of continuing medical education

*Mark only one oval per row.*

| | Often (at least 2-3 times per year) | Seldom (once per year) | Never |
|---|---|---|---|
| Training (in person) - rounds, in service, seminars | | | |
| Training (online) | | | |
| Local Conferences / meetings | | | |
| National Conferences/ Meetings | | | |
| Regional Conferences / Meetings - Within Africa | | | |
| International Conferences / Meetings | | | |

25. How often do your Radiographer/Technologists attend the following forms of continuing medical education

*Mark only one oval per row.*

| | Often (at least 2-3 times per year) | Seldom (once per year) | Never |
|---|---|---|---|
| Training (in person) - rounds, in service, seminars | ◯ | ◯ | ◯ |
| Training (online) | ◯ | ◯ | ◯ |
| Local Conferences / meetings | ◯ | ◯ | ◯ |
| National Conferences/ Meetings | ◯ | ◯ | ◯ |
| Regional Conferences / Meetings - Within Africa | ◯ | ◯ | ◯ |
| International Conferences / Meetings | ◯ | ◯ | ◯ |

26. Would your staff be interested in attending a Local/National/Regional MRI Workshop?

*Mark only one oval.*

◯ Yes

◯ No

27. Please could you elaborate on specific reasons for your selection below. Is access (including but not limited to costs) to training opportunities a challenge?

#### Health Economics

28. What is the average cost of an MRI scan at your facility (in US Dollars) *

_______________________

29. How is the cost of MRI scans covered *

*Check all that apply.*

- [ ] Fully public funded - National Health Insurance Scheme
- [ ] Some public funding
- [ ] Private (out-of-pocket)
- [ ] Private Insurance
- [ ] Hospital/clinic-funded

Other: [ ] _______________________

#### Research

30. What areas of MRI research has your facility participated in or interested in participating in. Please select all that apply. *

*Check all that apply.*

- [ ] Physics- hardware (scanner development, coil development)
- [ ] Physics - Pulse sequence development
- [ ] Image Processing
- [ ] Brain Imaging Application
- [ ] Cardiac Imaging Application
- [ ] Tumour Imaging Application
- [ ] Musculoskeletal Imaging Application
- [ ] Neuroscience
- [ ] Informatics and Radiomics

31. Does your facility lead or participate in research using MRI *

*Mark only one oval.*

- Yes
- No

32. Does your facility have a Research Ethics Board or Institutional Research Board? *

*Mark only one oval.*

- Yes
- No

#### Optional - Challenges

The following questions are optional but provides an opportunity for you to describe in your own words some of the challenges you face at your facility with operating your MRI facility and with participating in MRI research.

We encourage you to please take some time to briefly describe your challenges to help CAMERA better understand how to create a sustainable framework to increase MRI access, research and use in Africa.

33. Please describe briefly some challenges faced by your facility that impairs its day-to-day operation of the MRI program?

34. Please describe briefly some challenges faced by your facility that impairs its ability to conduct or participate in MRI-related research?

35. Please describe briefly any challenges in personnel capacity that impairs your facility's ability to perform clinical MRI imaging well

36. Please describe any solution you envision for tackling challenges you face in using MRI clinically

37. Please describe any solution you envision for tackling challenges you face in conducting MRI research

#### Optional - Contact information

The following questions are optional. Please complete these questions, if your facility wish to be included in future CAMERA initiatives, such as upcoming educational training opportunities, information for research collaborations, or be part of the CAMERA Research Network Database.

38. Please provide the name of your of facility

39. Please provide the address of your facility - Street number, name, city, province/state, country

40. Postal Code

41. Name of the primary contact person at your facility (Chief Radiographer or Chief Radiologist, or Managing Director)

42. Title/Role of the primary contact person

43. Contact phone number of the primary contact person

44. Email address of the primary contact person

Disclaimer

This survey tool is designed by the CAMERA MRI Environmental Scan Task Force comprising of the following individuals; Udunna Anazodo PhD, Godwin Ogbole MBBS, Edward Chege Nganga MBBS , Henk-Jan Mustaerts MD PhD, Mario Forjaz Secca PhD, and Johnes Obungoloch, PhD.

Information captured in this survey is confidential and will be solely accessible to the members of the CAMERA MRI Environmental Scan Task Force listed above. The information captured in this survey will be analyzed and maybe be published as part of a larger effort to describe the state of MRI access, use and readiness for research in Africa.

This survey tool has modified some questions from RAD-AID Radiology-Readiness Tool. For more information on RAD-AID Radiology-Readiness Tool please see https://www.rad-aid.org/resource-center/radiology-readiness

This content is neither created nor endorsed by Google.

Google Forms
